# Waning of BNT162b2 vaccine protection against SARS-CoV-2 infection in Qatar

**DOI:** 10.1101/2021.08.25.21262584

**Authors:** Hiam Chemaitelly, Patrick Tang, Mohammad R. Hasan, Sawsan AlMukdad, Hadi M. Yassine, Fatiha M. Benslimane, Hebah A. Al Khatib, Peter Coyle, Houssein H. Ayoub, Zaina Al Kanaani, Einas Al Kuwari, Andrew Jeremijenko, Anvar Hassan Kaleeckal, Ali Nizar Latif, Riyazuddin Mohammad Shaik, Hanan F. Abdul Rahim, Gheyath K. Nasrallah, Mohamed Ghaith Al Kuwari, Hamad Eid Al Romaihi, Adeel A. Butt, Mohamed H. Al-Thani, Abdullatif Al Khal, Roberto Bertollini, Laith J. Abu-Raddad

## Abstract

**BACKGROUND:** Waning of vaccine protection against SARS-CoV-2 infection or COVID-19 disease is a concern. This study investigated persistence of BNT162b2 (Pfizer-BioNTech) vaccine effectiveness against infection and disease in Qatar, where the Beta and Delta variants have dominated incidence and PCR testing is done at a mass scale.

**METHODS:** A matched test-negative, case-control study design was used to estimate vaccine effectiveness against SARS-CoV-2 infection and against any severe, critical, or fatal COVID-19 disease, between January 1, 2021 to August 15, 2021.

**RESULTS:** Estimated BNT162b2 effectiveness against any infection, asymptomatic or symptomatic, was negligible for the first two weeks after the first dose, increased to 36.5% (95% CI: 33.1-39.8) in the third week after the first dose, and reached its peak at 72.1% (95% CI: 70.9-73.2) in the first five weeks after the second dose. Effectiveness declined gradually thereafter, with the decline accelerating ≥15 weeks after the second dose, reaching diminished levels of protection by the 20^th^ week. Effectiveness against symptomatic infection was higher than against asymptomatic infection, but still waned in the same fashion. Effectiveness against any severe, critical, or fatal disease increased rapidly to 67.7% (95% CI: 59.1-74.7) by the third week after the first dose, and reached 95.4% (95% CI: 93.4-96.9) in the first five weeks after the second dose, where it persisted at about this level for six months.

**CONCLUSIONS:** BNT162b2-induced protection against infection appears to wane rapidly after its peak right after the second dose, but it persists at a robust level against hospitalization and death for at least six months following the second dose.

## Introduction

Qatar launched a mass Coronavirus Disease 2019 (COVID-19) immunization campaign on December 21, 2020, first using the BNT162b2^1^ (Pfizer-BioNTech) mRNA vaccine,^2^ and three months later adding the mRNA-1273^3^ (Moderna) vaccine.^4^ Immunization with both vaccines followed the FDA-approved protocol,^1, 3^ and vaccine coverage increased steadily from December 2020 until the present (Figure 1A). Vaccine rollout proceeded in phases in which vaccination was prioritized first to frontline healthcare workers, persons with severe or multiple chronic conditions, and persons ≥70 years of age. Age was the principal criterion for vaccine eligibility throughout the rollout. As of August 22, 2021, it is estimated that just over 80% of persons ≥12 years of age have received both doses of these mRNA vaccines.^5^ This appears to be the highest mRNA vaccine coverage worldwide.^6^

**Figure 1.**
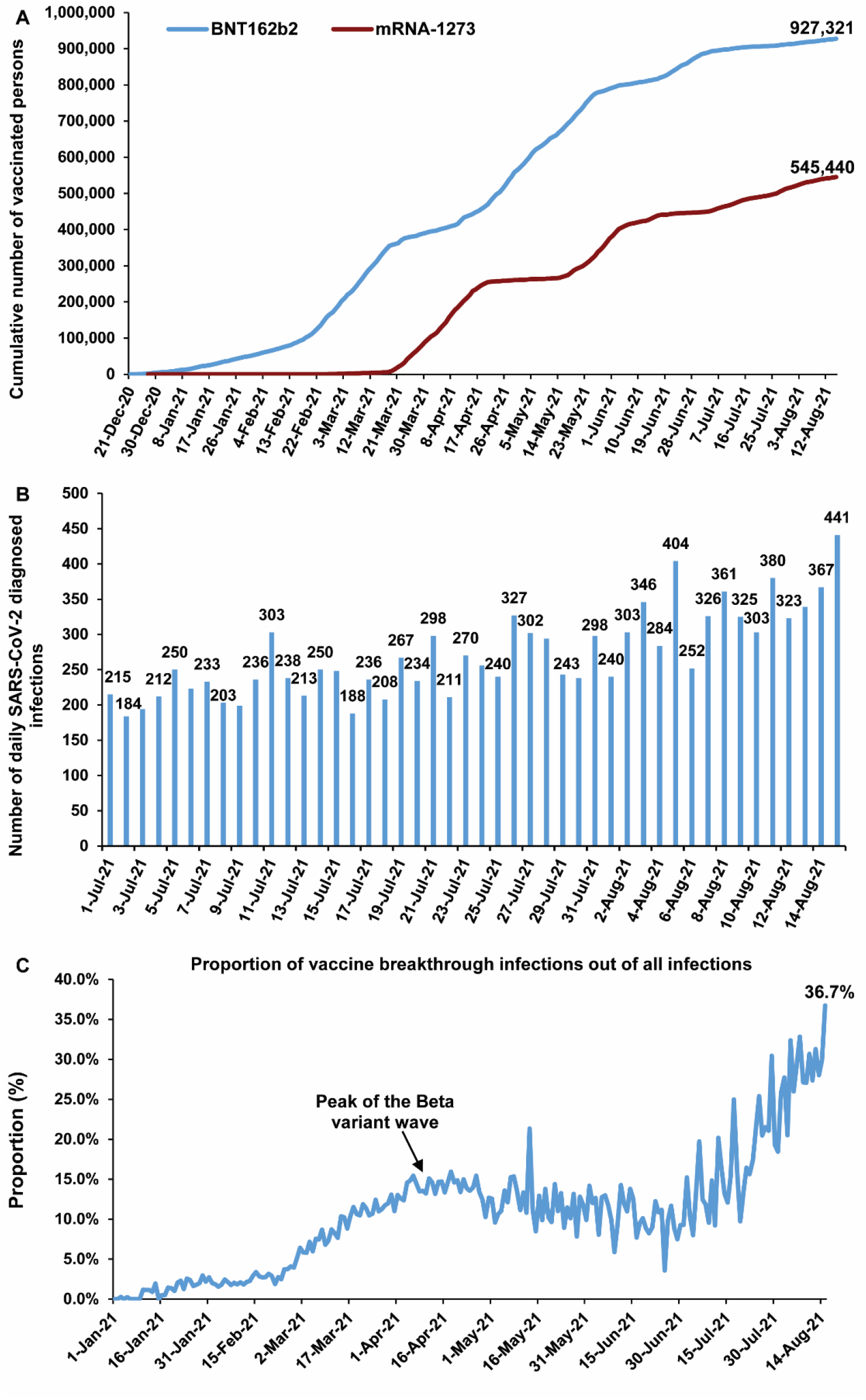
Time trend of A) cumulative numbers of BNT162b2-vaccinated and mRNA-1273-vaccinated persons, B) numbers of daily diagnosed SARS-CoV-2 infections in recent weeks, and C) proportion of vaccine breakthrough infections out of all infections, in Qatar.

As vaccination was scaled up, the country experienced two back-to-back severe acute respiratory syndrome coronavirus 2 (SARS-CoV-2) waves from January-June, 2021, which were dominated by the Alpha^7^ (B.1.1.7) and Beta^7^ (B.1.351) variants.^2, 4, 8–10^ Appreciable community transmission of the Delta^7^ (B.1.617.2) variant was first detected toward the end of March, 2021, and by August 22, 2021, Delta became the dominant variant.^8–10^ Despite the high vaccine coverage, the incidence of SARS-CoV-2 infection has been slowly increasing in recent weeks, although it remains at a relatively low level (Figure 1B).

In this study, we assessed the real-world effectiveness of the BNT162b2 vaccine over time after receiving the first and second doses, against SARS-CoV-2 infection and against COVID-19 hospitalization and death.

## Methods

### Study population, data sources, and study design

This study was conducted in the resident population of Qatar. COVID-19 laboratory testing, vaccination, clinical infection data, and related demographic details were extracted from the integrated, nationwide, digital-health information platform that hosts the national, federated SARS-CoV-2 databases. These databases are complete, and have captured all SARS-CoV-2-related data and demographic details with no missing information since the start of the epidemic. These include all records of polymerase chain reaction (PCR) testing, vaccinations, and COVID-19 hospitalizations.

Every PCR test conducted in Qatar, regardless of location or setting, is classified on the basis of symptoms and the reason for testing (clinical symptoms, contact tracing, random testing campaigns (surveys), individual requests, routine healthcare testing, pre-travel, and at port of entry). Qatar has unusually young, diverse demographics, in that only 9% of its residents are ≥50 years of age, and 89% are expatriates from over 150 countries.^11, 12^

Nearly all individuals in the population were vaccinated free of charge in Qatar, rather than elsewhere. In rare situations in which an individual was vaccinated outside Qatar, that individual’s vaccination details were still recorded in the health system at the port of entry upon return to Qatar, following national requirements and to benefit from privileges associated with vaccination, such as exemption from quarantine.^13^

Vaccine effectiveness against SARS-CoV-2 infection was estimated using the test-negative, case-control study design, a standard design for assessing vaccine effectiveness against influenza^14, 15^ and SARS-CoV-2.^2, 4, 14–18^ Key to this design is its control of bias arising from misclassification of infection and differences in health care-seeking behavior between vaccinated and unvaccinated individuals.^14, 15^ Cases (PCR-positive persons) and controls (PCR-negative persons) were matched one-to-one by sex, 10-year age group, nationality, reason for SARS-CoV-2 PCR testing, and calendar week of PCR test. Matching of cases and controls was performed to control for known differences in the risk of exposure to SARS-CoV-2 infection in Qatar^11, 19–21^

Only the first PCR-positive test during the study, January 1, 2021 to August 15, 2021, was included for each case, and only the first PCR-negative test during the study was included for each control. All PCR-negative tests for persons included as cases were excluded from analysis; that is no person was included as both a case and a control. These inclusion and exclusion criteria were implemented to control potential bias arising from repeated testing, such as a PCR-positive person undergoing a second PCR test a few days after infection diagnosis to test for clearance of infection. Modifications to these inclusion and exclusion criteria were investigated in sensitivity analyses as described below.

Effectiveness was estimated against documented infection (defined as a PCR-positive swab, regardless of the reason for PCR testing or the presence of symptoms), as well as against any severe,^22^ critical,^22^ or fatal^23^ COVID-19 disease. Classification of COVID-19 case severity (acute-care hospitalizations),^22^ criticality (ICU hospitalizations),^22^ and fatality^23^ followed World Health Organization (WHO) guidelines, and assessments were made by trained medical personnel using individual chart reviews. Each person who had a positive PCR test result and hospital admission was subject to an infection severity assessment every three days until discharge or death. Individuals who progressed to COVID-19 disease between the time of the PCR-positive test result and the end of the study were classified based on their worst outcome, starting with death,^23^ followed by critical disease,^22^ and then severe disease.^22^ Details of the COVID-19 severity, criticality, and fatality classification are found in Supplementary Section 1.

All records of PCR testing for those vaccinated and unvaccinated during the study were examined. All persons who received mixed vaccines, or who received a vaccine other than BNT162b2 were excluded. Every case that met the inclusion criteria and that could be matched to a control was included in the analysis. Both PCR-test outcomes and vaccination status were ascertained at the time of the PCR test.

The study was approved by the Hamad Medical Corporation and Weill Cornell Medicine-Qatar Institutional Review Boards with waiver of informed consent. Reporting of the study followed STROBE guidelines (Supplementary Table 1).

### Laboratory methods and classification of infections by variant type

Details of laboratory methods for real-time reverse-transcription PCR (RT-qPCR) testing are found in Supplementary Section 2. Methods for classification of infections by variant type using RT-qPCR variant screening^24^ of random positive clinical samples^8, 10^ are included in Supplementary Section 3. All PCR testing was conducted at the Hamad Medical Corporation Central Laboratory or at Sidra Medicine Laboratory, following standardized protocols.

### Statistical analysis

All records of PCR testing in Qatar during the study were examined in this study, but only samples of matched cases and controls were included in the analysis. Socio-demographic characteristics of study samples were described using frequency distributions and measures of central tendency. Differences in proportions across categorical variables between study groups were evaluated using Chi-square tests. A two-sided p-value of <0.05 indicated a significant association.

The odds ratio, comparing odds of vaccination among cases versus controls, and its associated 95% confidence interval (CI) were calculated using the exact method, or the Cornfield method, when no vaccinations were recorded among cases or controls. Confidence intervals were not adjusted for multiplicity. Interactions were not investigated. Vaccine effectiveness at different time points and its associated 95% CI were then calculated by applying the following equation:^14, 15^

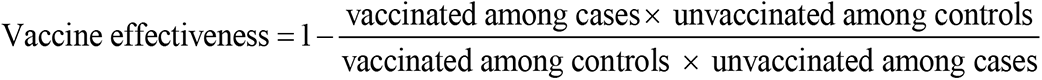

To ensure that vaccine effectiveness estimates were not biased by epidemic phase,^14, 25^ the gradual roll-out of vaccination during the study,^14, 25^ or other confounders,^26, 27^ a sensitivity analysis was conducted by adjusting for prior infection and matching factors in logistic regression, that is, by sex, age, nationality, reason for PCR testing, and calendar week of PCR test.

Vaccine effectiveness was also estimated against symptomatic infection, defined as a PCR-positive test conducted because of clinical suspicion due to presence of symptoms compatible with a respiratory tract infection, and against asymptomatic infection, defined as a PCR-positive test conducted with no reported presence of symptoms compatible with a respiratory tract infection. In the latter case, PCR testing was done strictly as part of a survey, for pre-travel requirement, or at port of entry into the country. Vaccine effectiveness was further estimated in subgroup analyses stratifying cases and controls by age, variant type, or severe forms of COVID-19 disease.

Several additional sensitivity analyses were conducted by modifying the study inclusion and exclusion criteria or by incorporating an additional matching factor to investigate whether the effectiveness estimates could have been biased by an unknown factor. Descriptions of these sensitivity analyses can be found in Supplementary Section 4.

To provide further validation of study results, effectiveness was further estimated by deriving adjusted odds ratios (AORs) from multivariable logistic regression analyses of associations with a PCR-positive test, using the full unmatched sample, that is, by applying a different method from that of the main analysis of matched test-negative, case-control study design. Statistical analyses were conducted in STATA/SE version 17.0.^28^

## Results

### Study population

Between December 21, 2020 and August 15, 2021, 927,321 individuals received at least one dose of BNT162b2, and 891,481 completed the two-dose regimen (Figure 1A). The median date at first dose was April 20, 2021, and the median date at second dose was May 9, 2021. The median time elapsed between the first and second doses was 21 days (interquartile range (IQR): 21-22 days), and 97.5% of individuals received their second dose ≤30 days after the first dose. Notably, during this period, 545,440 individuals received at least one dose of mRNA-1273, and 451,732 completed the two-dose regimen (Figure 1A).

Supplementary Figure 1 presents a flowchart describing the population selection process for investigating and estimating BNT162b2 effectiveness against SARS-CoV-2 infection. Demographic characteristics and reasons for PCR testing of samples used to estimate vaccine effectiveness are presented in Table 1. The median age of study subjects was 31-32 years, 70.6% were males, and subjects came from diverse national origins. Study samples were representative of the unique demographics of the population of Qatar.^11, 12^

**Table 1.** Demographic characteristics of subjects and reasons for PCR testing among samples used to estimate BNT162b2 vaccine effectiveness. The table strictly includes samples used in the 0-4-weeks-after-second-dose analysis. However, samples used in the remaining analyses for other time intervals are similar, as they differ only in the inclusion of the specific vaccination subpopulation category (such as persons at 5-9 weeks after the second dose).

| Characteristics | Cases*<br>(PCR-positive) | Controls*<br>(PCR-negative) | p-value |
| --- | --- | --- | --- |
| **Median age (IQR) — years** | 32 (23-39) | 31 (23-39) | 0.410 |
| **Age group — no. (%)** |  |  |  |
| <20 years | 27,890 (19.4) | 27,890 (19.4) | 1.000 |
| 20-29 years | 34,716 (24.1) | 34,716 (24.1) |  |
| 30-39 years | 48,770 (33.9) | 48,770 (33.9) |  |
| 40-49 years | 23,607 (16.4) | 23,607 (16.4) |  |
| 50-59 years | 7,255 (5.0) | 7,255 (5.0) |  |
| 60-69 years | 1,458 (1.0) | 1,458 (1.0) |  |
| 70+ years | 379 (0.3) | 379 (0.3) |  |
| **Sex** |  |  |  |
| Male | 101,766 (70.6) | 101,766 (70.6) | 1.000 |
| Female | 42,309 (29.4) | 42,309 (29.4) |  |
| **Nationality<sup>†</sup>** |  |  |  |
| Bangladeshi | 10,381 (7.2) | 10,381 (7.2) | 1.000 |
| Egyptian | 7,404 (5.1) | 7,404 (5.1) |  |
| Filipino | 14,453 (10.0) | 14,453 (10.0) |  |
| Indian | 43,796 (30.4) | 43,796 (30.4) |  |
| Nepalese | 12,914 (9.0) | 12,914 (9.0) |  |
| Pakistani | 7,431 (5.2) | 7,431 (5.2) |  |
| Qatari | 17,847 (12.4) | 17,847 (12.4) |  |
| Sri Lankan | 4,320 (3.0) | 4,320 (3.0) |  |
| Sudanese | 3,777 (2.6) | 3,777 (2.6) |  |
| Other nationalities<sup>‡</sup> | 21,752 (15.1) | 21,752 (15.1) |  |
| **Reason for PCR testing** |  |  |  |
| Clinical suspicion | 37,272 (25.9) | 37,272 (25.9) | 1.000 |
| Contact tracing | 17,946 (12.5) | 17,946 (12.5) |  |
| Healthcare routine testing | 14,151 (9.8) | 14,151 (9.8) |  |
| Survey | 26,945 (18.7) | 26,945 (18.7) |  |
| Port of entry | 30,486 (21.2) | 30,486 (21.2) |  |
| Pre-travel | 4,539 (3.2) | 4,539 (3.2) |  |
| Individual request | 12,476 (8.7) | 12,476 (8.7) |  |
| Other | 260 (0.2) | 260 (0.2) |  |

Abbreviations: IQR, interquartile range; PCR, polymerase chain reaction.
\*Cases and controls were matched one-to-one by sex, 10-year age group, nationality, reason for PCR testing, and calendar week of PCR test.
<sup>†</sup>Nationalities were chosen to represent the most populous groups in Qatar.
<sup>‡</sup>These comprise 118 other nationalities in Qatar.

Only 25.9% of cases (PCR-confirmed infections) were diagnosed because of symptoms (Table 1). The remaining cases were diagnosed because of PCR testing for other reasons, including contact tracing, random testing campaigns (surveys), individual requests, routine healthcare testing, pre-travel, and at port of entry.

### Vaccine breakthrough infections

As of the end of the study, August 15, 2021, 8,155 and 8,935 SARS-CoV-2 breakthrough infections had been recorded among those who received either one or two doses of BNT162b2, respectively. The percentage of vaccine (BNT162b2 or mRNA-1273) breakthrough infections out of the daily diagnosed infections increased gradually with time and was at 36.7% on August 15, 2021 (Figure 1C). Most vaccine breakthrough infections (76.9%) were recorded for the BNT162b2 vaccine.

Also, as of August 15, 2021, 377 and 96 severe COVID-19 disease cases (acute-care hospitalizations;^22^ Supplementary Section 1) had been recorded among those who received either one or two doses of BNT162b2, respectively. Similarly, 31 and 8 critical COVID-19 disease cases (ICU-care hospitalizations;^22^ Supplementary Section 1), and 34 and 15 fatal COVID-19 disease cases (COVID-19 deaths;^23^ Supplementary Section 1) had been also recorded, respectively.

### Vaccine effectiveness against any SARS-CoV-2 infection

Estimated BNT162b2 effectiveness against any SARS-CoV-2 infection was negligible for the first two weeks after the first dose, increased to 36.5% (95% CI: 33.1-39.8) in the third week after the first dose, and reached its peak at 72.1% (95% CI: 70.9-73.2) in the first five weeks after the second dose (Table 2 and Figure 2). However, effectiveness declined gradually starting from 0-4 weeks after the second dose, and the decline accelerated ≥15 weeks after the second dose. Effectiveness was diminished ≥20 weeks after the second dose.

**Figure 2.**
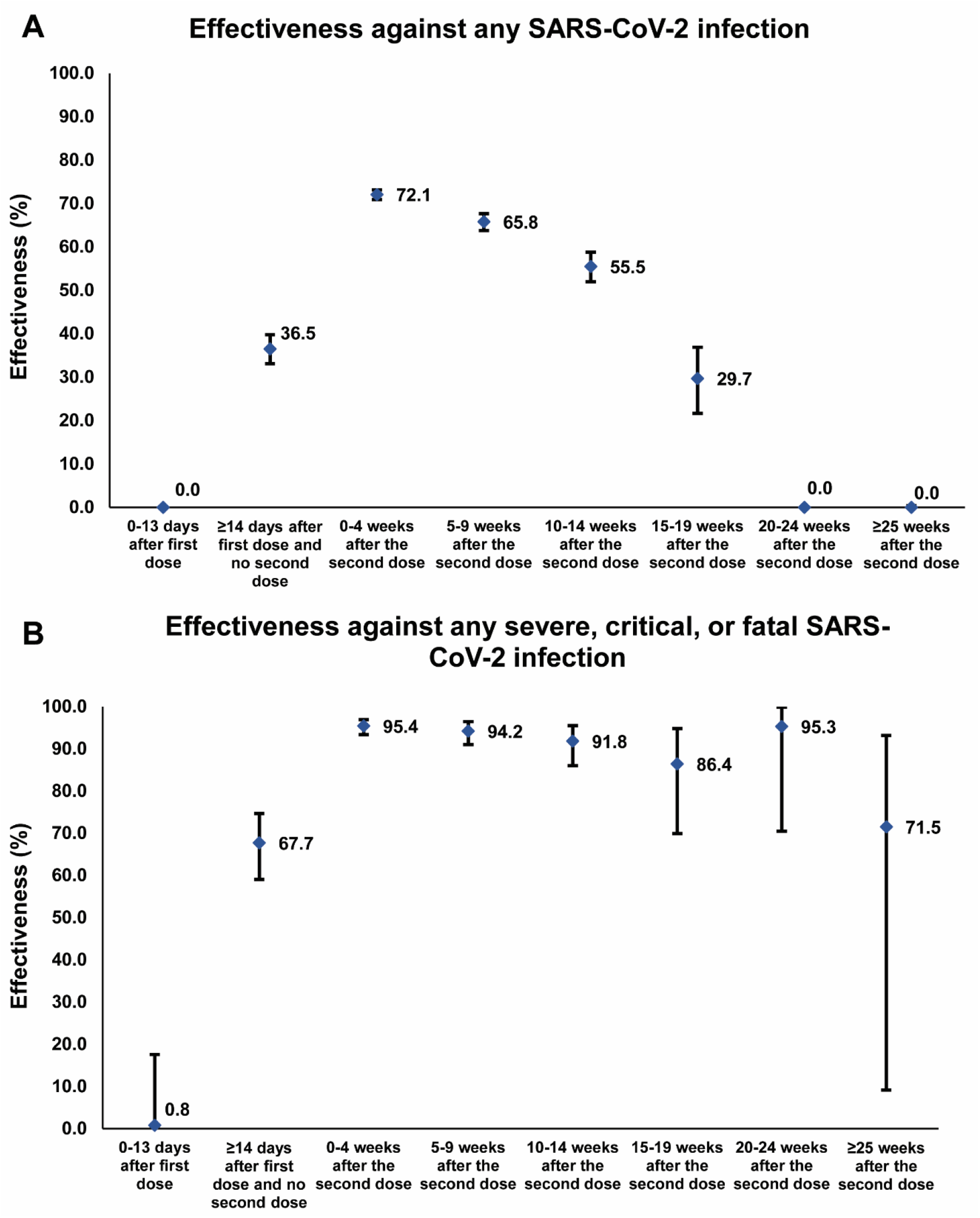
Effectiveness of the BNT162b2 vaccine against A) any SARS-CoV-2 infection and B) any severe, critical, or fatal COVID-19 disease. Data are presented as effectiveness point estimates with error bars indicating the corresponding 95% confidence intervals.

**Table 2.** Effectiveness of the BNT162b2 vaccine against any SARS-CoV-2 infection and against any severe, critical, or fatal COVID-19 disease.

|  | Effectiveness against infection |  |  |  |  | Effectiveness against hospitalization and death |  |  |  |  |
| --- | --- | --- | --- | --- | --- | --- | --- | --- | --- | --- |
|  | Cases*<br>(PCR-positive) |  | Controls*<br>(PCR-negative) |  | Effectiveness<br>in %<br>(95% CI)<sup>†</sup> | Cases*<br>(Severe, critical, or fatal disease)<sup>‡</sup> |  | Controls*<br>(PCR-negative) |  | Effectiveness<br>in %<br>(95% CI)<sup>†</sup> |
|  | Vaccinated | Unvaccinated | Vaccinated | Unvaccinated |  | Vaccinated | Unvaccinated | Vaccinated | Unvaccinated |  |
| 0-13 days after first dose | 4,194 | 138,900 | 3,976 | 139,118 | 0.0<br>(0.0-0.0) | 249 | 3,982 | 251 | 3,980 | 0.8<br>(0.0-17.6) |
| ≥14 days after first dose and no second dose | 2,345 | 139,349 | 3,659 | 138,035 | 36.5<br>(33.1-39.8) | 98 | 4,017 | 289 | 3,826 | 67.7<br>(59.1-74.7) |
| 0-4 weeks after the second dose | 3,141 | 140,934 | 10,659 | 133,416 | 72.1<br>(70.9-73.2) | 32 | 4,078 | 598 | 3,512 | 95.4<br>(93.4-96.9) |
| 5-9 weeks after the second dose | 1,612 | 139,694 | 4,610 | 136,696 | 65.8<br>(63.8-67.7) | 21 | 4,050 | 334 | 3,737 | 94.2<br>(91.0-96.5) |
| 10-14 weeks after the second dose | 1,006 | 139,230 | 2,242 | 137,994 | 55.5<br>(52.0-58.8) | 15 | 4,000 | 175 | 3,840 | 91.8<br>(86.0-95.5) |
| 15-19 weeks after the second dose | 581 | 138,828 | 825 | 138,584 | 29.7<br>(21.7-36.9) | 7 | 3,984 | 51 | 3,940 | 86.4<br>(69.9-94.8) |
| 20-24 weeks after the second dose | 608 | 138,676 | 523 | 138,761 | 0.0<br>(0.0-0.0) | 1 | 3,971 | 21 | 3,951 | 95.3<br>(70.5-99.9) |
| ≥25 weeks after the second dose | 483 | 138,702 | 425 | 138,760 | 0.0<br>(0.0-0.4) | 4 | 3,979 | 14 | 3,969 | 71.5<br>(9.2-93.2) |

Abbreviations: CI, confidence interval; PCR, polymerase chain reaction.
\*Cases and controls were matched one-to-one by sex, 10-year age group, nationality, reason for PCR testing, and calendar week of PCR test.
<sup>†</sup>Vaccine effectiveness was estimated using the test-negative, case-control study design.<sup>14,15</sup>
<sup>‡</sup>Severity,<sup>22</sup> criticality,<sup>22</sup> and fatality<sup>23</sup> were defined as per World Health Organization guidelines.

A sensitivity analysis adjusting for prior infection, sex, age, nationality, reason for PCR testing, and calendar week of PCR test in logistic regression, confirmed main analysis results (Table 3).

**Table 3.** Sensitivity analysis. Effectiveness of the BNT162b2 vaccine against any SARS-CoV-2 infection and against any severe, critical, or fatal COVID-19 disease, after adjusting for sex, age, nationality, reason for PCR testing, calendar week of PCR test, and prior infection.

|  | Effectiveness against symptomatic infection* |  |  |  |  | Effectiveness against asymptomatic infection† |  |  |  |  |
| --- | --- | --- | --- | --- | --- | --- | --- | --- | --- | --- |
|  | Cases‡<br>(PCR-positive) |  | Controls‡<br>(PCR-negative) |  | Effectiveness<br>in %<br>(95% CI)§ | Cases‡<br>(PCR-positive) |  | Controls‡<br>(PCR-negative) |  | Effectiveness<br>in %<br>(95% CI)§ |
|  | Vaccinated | Unvaccinated | Vaccinated | Unvaccinated |  | Vaccinated | Unvaccinated | Vaccinated | Vaccinated |  |
| 0-13 days after first dose | 1,833 | 35,028 | 1,764 | 35,097 | 0.0<br>(0.0-2.7) | 1,111 | 60,562 | 1,062 | 60,611 | 0.0<br>(0.0-3.9) |
| ≥14 days after first dose and no second dose | 976 | 35,453 | 1,812 | 34,617 | 47.4<br>(43.0-51.5) | 635 | 60,551 | 831 | 60,355 | 23.8<br>(15.4-31.4) |
| 0-4 weeks after the second dose | 749 | 36,523 | 3,412 | 33,860 | 79.6<br>(77.9-81.2) | 1,222 | 60,748 | 3,253 | 58,717 | 63.7<br>(61.2-66.1) |
| 5-9 weeks after the second dose | 537 | 35,619 | 1,776 | 34,380 | 70.8<br>(67.8-73.6) | 673 | 60,635 | 1,472 | 59,836 | 54.9<br>(50.5-58.9) |
| 10-14 weeks after the second dose | 342 | 35,303 | 856 | 34,789 | 60.6<br>(55.2-65.4) | 492 | 60,572 | 796 | 60,268 | 38.5<br>(31.0-45.2) |
| 15-19 weeks after the second dose | 168 | 35,015 | 332 | 34,851 | 49.6<br>(39.1-58.4) | 337 | 60,536 | 333 | 60,540 | 0.0<br>(0.0-13.3) |
| 20-24 weeks after the second dose | 172 | 34,933 | 171 | 34,934 | 0.0<br>(0.0-19.1) | 359 | 60,506 | 269 | 60,596 | 0.0<br>(0.0-0.0) |
| ≥25 weeks after the second dose | 148 | 34,946 | 126 | 34,968 | 0.0<br>(0.0-8.0) | 272 | 60,513 | 227 | 60,558 | 0.0<br>(0.0-0.0) |

Abbreviations: CI, confidence interval; PCR, polymerase chain reaction.
\*Cases and controls were matched one-to-one by sex, 10-year age group, nationality, reason for PCR testing, and calendar week of PCR test.

### Vaccine effectiveness against any SARS-CoV-2 infection by age and by variant

BNT162b2 effectiveness was assessed for those <60 years of age and those ≥60, to investigate whether declining effectiveness over time could have been confounded by age. Results for both age groups were similar, in scale (but slightly lower for those ≥60 years of age), in pattern of declining effectiveness (Supplementary Table 2), and to those for all subjects of all age groups (Table 2).

The above effectiveness measures largely reflect BNT162b2 effectiveness against Beta,^2, 29^ which was by far the dominant variant during most of these time periods.^2, 4, 8–10^ However, with the steady increase in incidence of Delta during the summer of 2021,^8–10, 30^ and the steady decline in Beta incidence during this time,^8–10^ effectiveness measures ≥15 weeks after the second dose increasingly reflected BNT162b2 effectiveness against Delta. Notably, although Qatar experienced an Alpha variant wave early in 2021, this wave peaked in the first week of March, 2021, and at a much lower incidence than the peak of the Beta variant wave, which occurred in April, 2021.^2, 4, 8–10^ Most incidence of Alpha occurred at a time when the number of vaccinated persons was still small; thus, Alpha infections did not contribute appreciably to these effectiveness measures.

BNT162b2 effectiveness was assessed against each of Alpha, Beta, and Delta infections to investigate whether the declining effectiveness could have been confounded by exposure to different variants over time. Estimated effectiveness against each of these variants (Supplementary Table 3) demonstrated a similar pattern to that seen against any SARS-CoV-2 infection (Table 2). However, estimates for individual variants had wider 95% confidence intervals, because they were derived using smaller numbers of confirmed PCR-positive cases, that is, only those confirmed as Alpha, Beta, or Delta using RT-qPCR genotyping (Supplementary Section 3). In the analysis of any SARS-CoV-2 infection (Table 2), the study sample included 209,875 cases (Supplementary Figure 1), while in the analyses of Alpha, Beta, and Delta infections, study samples included only 3,233, 5,480, and 3,223 cases, respectively, and these were diagnosed mostly during the summer of 2021 (Supplementary Section 3). Variant RT-qPCR genotyping started at a considerable scale only in the early summer of 2021, well after the large Beta wave had peaked in April, 2021, thus explaining the small samples sizes in these analyses.

### Vaccine effectiveness against symptomatic and asymptomatic infections

Estimated BNT162b2 effectiveness against each of symptomatic infection and asymptomatic infection demonstrated the same pattern of increasing effectiveness after the first dose, peak effectiveness in the first five weeks after the second dose, and a gradual decline in effectiveness in the following weeks that accelerated ≥20 weeks after the second dose for symptomatic infection, and ≥15 weeks after the second dose for asymptomatic infection (Table 4). However, effectiveness against symptomatic infection was consistently higher than that against asymptomatic infection. The peak effectiveness against symptomatic infection was 79.6% (95% CI: 77.9-81.2) while that against asymptomatic infection was only 63.7% (95% CI: 61.2-66.1).

**Table 4.** Effectiveness of the BNT162b2 vaccine against each of symptomatic SARS-CoV-2 infection and asymptomatic SARS-CoV-2 infection.

|  | Effectiveness against symptomatic infection* |  |  |  |  | Effectiveness against asymptomatic infection† |  |  |  |  |
| --- | --- | --- | --- | --- | --- | --- | --- | --- | --- | --- |
|  | Cases‡<br>(PCR-positive) |  | Controls‡<br>(PCR-negative) |  | Effectiveness<br>in %<br>(95% CI)§ | Cases‡<br>(PCR-positive) |  | Controls‡<br>(PCR-negative) |  | Effectiveness<br>in %<br>(95% CI)§ |
|  | Vaccinated | Unvaccinated | Vaccinated | Unvaccinated |  | Vaccinated | Unvaccinated | Vaccinated | Vaccinated |  |
| ≥14 days after first dose and no second dose | 976 | 35,453 | 1,812 | 34,617 | 47.4<br>(43.0-51.5) | 635 | 60,551 | 831 | 60,355 | 23.8<br>(15.4-31.4) |
| 0-4 weeks after the second dose | 749 | 36,523 | 3,412 | 33,860 | 79.6<br>(77.9-81.2) | 1,222 | 60,748 | 3,253 | 58,717 | 63.7<br>(61.2-66.1) |
| 5-9 weeks after the second dose | 537 | 35,619 | 1,776 | 34,380 | 70.8<br>(67.8-73.6) | 673 | 60,635 | 1,472 | 59,836 | 54.9<br>(50.5-58.9) |
| 10-14 weeks after the second dose | 342 | 35,303 | 856 | 34,789 | 60.6<br>(55.2-65.4) | 492 | 60,572 | 796 | 60,268 | 38.5<br>(31.0-45.2) |
| 15-19 weeks after the second dose | 168 | 35,015 | 332 | 34,851 | 49.6<br>(39.1-58.4) | 337 | 60,536 | 333 | 60,540 | 0.0<br>(0.0-13.3) |
| 20-24 weeks after the second dose | 172 | 34,933 | 171 | 34,934 | 0.0<br>(0.0-19.1) | 359 | 60,506 | 269 | 60,596 | 0.0<br>(0.0-0.0) |
| ≥25 weeks after the second dose | 148 | 34,946 | 126 | 34,968 | 0.0<br>(0.0-8.0) | 272 | 60,513 | 227 | 60,558 | 0.0<br>(0.0-0.0) |

Abbreviations: CI, confidence interval; PCR, polymerase chain reaction.
\*A symptomatic infection was defined as a PCR-positive test conducted because of clinical suspicion due to presence of symptoms compatible with a respiratory tract infection.
†An asymptomatic infection was defined as a PCR-positive test conducted with no reported presence of symptoms compatible with a respiratory tract infection, that is the PCR testing was done as part of a survey, for pre-travel requirement, or at port of entry into the country.
‡Cases and controls were matched one-to-one by sex, 10-year age group, nationality, reason for PCR testing, and calendar week of PCR test.
§Vaccine effectiveness was estimated using the test-negative, case-control study design.<sup>14,15</sup>

### Vaccine effectiveness against COVID-19 hospitalization and death

Estimated BNT162b2 effectiveness against any severe, critical, or fatal disease due to any SARS-CoV-2 infection, was negligible for the first two weeks after the first dose. It increased rapidly to 67.7% (95% CI: 59.1-74.7) in the third week after the first dose, and reached a peak of 95.4% (95% CI: 93.4-96.9) in the first five weeks after the second dose (Table 2 and Figure 2). Unlike effectiveness against infection, there was no evident decline in this effectiveness over time. However, at ≥25 weeks after the second dose, there was a hint of a decline in effectiveness, but the case numbers were small.

The sensitivity analysis adjusting for prior infection and the matching factors in logistic regression confirmed the main analysis results (Table 3). Effectiveness by age group (Supplementary Table 2) or by variant type (Supplementary Table 3) also showed similar results. BNT162b2 effectiveness was also estimated against each of severe disease, critical disease, and fatal disease (Supplementary Table 4), as opposed to against a composite outcome of any severe, critical, or fatal disease (Table 2). Estimated effectiveness against each of these individual disease outcomes was similar to that against the composite disease outcome, with no evident decline in effectiveness in the months following the second dose.

### Additional analyses

Six additional sensitivity analyses were conducted to investigate whether these real-world effectiveness estimates could have been biased by an unknown factor (Supplementary Section 4). All analyses generated consistent results indicating similar values for effectiveness measures, and the same pattern of declining effectiveness in the months following the second dose (Supplementary Tables 5-10), as observed in the main analysis (Table 2).

To further validate the study results, effectiveness was estimated using a different method from that of a matched test-negative, case-control study design. Estimates were derived using multivariable logistic regression analysis of associations with a PCR-positive test during the study, and adjusting for prior infection, sex, age, nationality, reason for PCR testing, and calendar week of PCR test (Supplementary Table 11). This analysis also generated consistent results, indicating similar values for effectiveness measures, and the same pattern of declining effectiveness in the months following the second dose, as in the main analysis (Table 2).

## Discussion

BNT162b2-induced protection against infection builds rapidly after the first dose, peaks in the first five weeks after the second dose, but then gradually wanes in subsequent months. The waning appears to accelerate ≥15 weeks after the second dose, with protection being reduced to a negligible level by the 20^th^ week. While the protection diminished faster against asymptomatic infection than against symptomatic infection, no evidence was found for any appreciable waning of protection against hospitalization and death, which remained robust at about 90% for six months following the second dose. Implications of these findings on infection transmission remain to be clarified, but vaccine breakthrough infections were found recently, in this same population, to be less infectious than primary infections in unvaccinated individuals.^31^

Since the immunization campaign prioritized vaccination of persons with severe or multiple chronic conditions and by age group, this pattern of waning of protection could theoretically be cofounded by effects of age and comorbidities. However, this possibility was not supported by our results, as the same pattern of waning of protection was observed for all ages. Notably, old age may serve as a proxy for co-morbid conditions and the number of persons with severe or multiple chronic conditions is small among the young, working age population of Qatar.^11, 12^

Infection incidence was driven by different variants over time; thus, it is possible that waning of protection could be confounded by exposure to different variants at different time points. However, this seems unlikely. By far the dominant variant during the study was Beta,^2, 4, 8–10^ and a similar pattern of waning of protection was observed for Alpha, Beta, and Delta.

Vaccinated persons presumably have a higher social contact rate than unvaccinated persons, and may also have reduced adherence to safety measures.^32–34^ This behavior could reduce real-world effectiveness of the vaccine compared to its biological effectiveness, possibly explaining the waning of protection. However, risk compensation is perhaps more likely to affect the overall level of effectiveness, rather than the waning of protection over time, unless such risk compensation increases with time after the second dose.

Emerging evidence supports the findings of this study. An increasing number of studies suggests significant waning of BNT162b2 effectiveness.^35–38^ The findings are also supported by recent reports from Israel and the United States (U.S.) indicating declining BNT162b2 effectiveness against infection with elapsed time and by calendar month.^39–42^ Our findings may also explain the observed low effectiveness against Delta in countries where the second dose was implemented three weeks after the first dose, such as in Israel,^39^ Qatar,^30^ and the U.S.,^42^ but higher effectiveness against Delta in countries where a delayed schedule has been implemented, such as in Canada^18^ and the United Kingdom.^16, 17^

This study has limitations. Data on co-morbid conditions were not available; therefore, they could not be explicitly factored into our analysis. However, adjusting for age may have served as a proxy given that co-morbidities are associated with older age. With the young population of Qatar,^11, 12^ only a small proportion of the study population may have had serious co-morbid conditions. Our findings may not be generalizable to other countries where the elderly constitute a sizable proportion of the total population.

Effectiveness was assessed using an observational, test-negative, case-control study design,^14, 15^ rather than a randomized, clinical trial design, in which cohorts of vaccinated and unvaccinated individuals were followed up. We have been unable to use a cohort study design due to depletion of the unvaccinated cohorts with the high vaccine coverage. However, the cohort study design applied earlier to the same population of Qatar, yielded similar findings to the test-negative case-control design,^2, 4^ supporting the validity of this standard approach in assessing vaccine effectiveness for respiratory tract infections.^2, 4, 14–18^ The results of this study are also consistent with our earlier effectiveness estimates immediately after the first and second doses,^2, 29^ noting that the earlier estimates were against (mostly) symptomatic infections with low PCR cycle threshold values, while the present study estimates are against (mostly) asymptomatic infections of both high and low PCR cycle threshold values.

Nonetheless, one cannot theoretically exclude the possibility that in real-world data, bias could arise in unexpected ways, or from unknown sources, such as subtle differences in test-seeking behavior or changes in the pattern of testing with introduction of other testing modalities, such as rapid antigen testing. However, the same findings were reached regardless of the reason for PCR testing, and regardless of whether it was following appearance of symptoms, or because of a mandatory requirement including routine healthcare testing, pre-travel, and at the port of entry. Moreover, with the mass scale of PCR testing in Qatar, where about 5% of the population are tested every week,^5^ the likelihood of bias is perhaps minimized. Indeed, the multiple sensitivity and additional analyses conducted to investigate bias, such as by modifying the inclusion and exclusion criteria in various ways, all presented consistent findings.

In conclusion, BNT162b2-induced protection against infection appears to peak in the first five weeks after the second dose, but then gradually wanes month by month, before reaching diminished levels by the 20^th^ week. Meanwhile, BNT162b2-induced protection against hospitalization and death appears to persist with hardly any waning for at least six months following the second dose. These findings suggest that a large proportion of the vaccinated population could lose its protection against infection in the coming months, perhaps increasing the potential for new epidemic waves. These findings argue for a booster vaccination to reinforce immunity against infection, and to avert possible waning in protection against hospitalization and death over time.

## Data Availability

The dataset of this study is a property of the Qatar Ministry of Public Health that was provided to the researchers through a restricted-access agreement that prevents sharing the dataset with a third party or publicly. Future access to this dataset can be considered through a direct application for data access to Her Excellency the Minister of Public Health (https://www.moph.gov.qa/english/Pages/default.aspx). Aggregate data are available within the manuscript and its Supplementary information.

## Acknowledgements

We acknowledge the many dedicated individuals at Hamad Medical Corporation, the Ministry of Public Health, the Primary Health Care Corporation, Qatar Biobank, Sidra Medicine, and Weill Cornell Medicine for their diligent efforts and contributions to make this study possible. The authors are grateful for institutional salary support from the Biomedical Research Program and the Biostatistics, Epidemiology, and Biomathematics Research Core, both at Weill Cornell Medicine-Qatar, as well as for institutional salary support provided by the Ministry of Public Health and Hamad Medical Corporation. The authors are also grateful for the Qatar Genome Programme for institutional support for the reagents needed for the viral genome sequencing. The funders of the study had no role in study design, data collection, data analysis, data interpretation, or writing of the article. Statements made herein are solely the responsibility of the authors.

## Author contributions

HC co-designed the study, performed the statistical analyses, and co-wrote the first draft of the article. LJA conceived and co-designed the study, led the statistical analyses, and co-wrote the first draft of the article. PT and MRH conducted the multiplex, RT-qPCR variant screening and viral genome sequencing. HY, FMB, and HAK conducted viral genome sequencing. All authors contributed to data collection and acquisition, database development, discussion and interpretation of the results, and to the writing of the manuscript. All authors have read and approved the final manuscript.

## Competing interests

Dr. Butt has received institutional grant funding from Gilead Sciences unrelated to the work presented in this paper. Otherwise, we declare no competing interests.

## Supplementary Appendix

### Supplementary Section 1. COVID-19 severity, criticality, and fatality classification

Severe Coronavirus Disease 2019 (COVID-19) disease was defined per the World health Organization (WHO) classification as a severe acute respiratory syndrome coronavirus 2 (SARS-CoV-2) infected person with “oxygen saturation of <90% on room air, and/or respiratory rate of >30 breaths/minute in adults and children >5 years old (or ≥60 breaths/minute in children <2 months old or ≥50 breaths/minute in children 2–11 months old or ≥40 breaths/minute in children 1–5 years old), and/or signs of severe respiratory distress (accessory muscle use and inability to complete full sentences, and, in children, very severe chest wall indrawing, grunting, central cyanosis, or presence of any other general danger signs)”.^1^ Detailed WHO criteria for classifying SARS-CoV-2 infection severity can be found in the WHO technical report.^1^

Critical COVID-19 disease was defined per WHO classification as a SARS-CoV-2 infected person with “acute respiratory distress syndrome, sepsis, septic shock, or other conditions that would normally require the provision of life sustaining therapies such as mechanical ventilation (invasive or non-invasive) or vasopressor therapy”.^1^ Detailed WHO criteria for classifying SARS-CoV-2 infection criticality can be found in the WHO technical report.^1^

COVID-19 death was defined per WHO classification as “a death resulting from a clinically compatible illness, in a probable or confirmed COVID-19 case, unless there is a clear alternative cause of death that cannot be related to COVID-19 disease (e.g. trauma). There should be no period of complete recovery from COVID-19 between illness and death. A death due to COVID-19 may not be attributed to another disease (e.g. cancer) and should be counted independently of preexisting conditions that are suspected of triggering a severe course of COVID-19”. Detailed WHO criteria for classifying COVID-19 death can be found in the WHO technical report.^2^

### Supplementary Section 2. Laboratory methods

Nasopharyngeal and/or oropharyngeal swabs were collected for PCR testing and placed in Universal Transport Medium (UTM). Aliquots of UTM were: extracted on a QIAsymphony platform (QIAGEN, USA) and tested with real-time reverse-transcription PCR (RT-qPCR) using TaqPath™ COVID-19 Combo Kits (Thermo Fisher Scientific, USA) on an ABI 7500 FAST (Thermo Fisher, USA); tested directly on the Cepheid GeneXpert system using the Xpert Xpress SARS-CoV-2 (Cepheid, USA); or loaded directly into a Roche cobas® 6800 system and assayed with a cobas® SARS-CoV-2 Test (Roche, Switzerland). The first assay targets the viral S, N, and ORF1ab gene regions. The second targets the viral N and E-gene regions, and the third targets the ORF1ab and E-gene regions.

All PCR testing was conducted at the Hamad Medical Corporation Central Laboratory or Sidra Medicine Laboratory, following standardized protocols.

### Supplementary Section 3. Classification of infections by variant type

Surveillance for SARS-CoV-2 variants in Qatar is based on viral genome sequencing and multiplex, real-time reverse-transcription PCR (RT-qPCR) variant screening^3^ of random positive clinical samples,^4–8^ and complemented by deep sequencing of wastewater samples.^6^ The ascertainment of the B.1.1.7 (Alpha^9^), B.1.351 (Beta^9^), and B.1.617.2 (Delta^9^) cases in this study was based on the results of weekly RT-qPCR genotyping of the positive clinical samples.^6, 8^ Between March 22, 2021 and August 3, 2021, RT-qPCR genotyping identified 5,480 (45.8%) B.1.351-like cases, 3,233 (27.0%) B.1.1.7-like cases, 3,223 (26.9%) “other” variant cases, and 41 (0.3%) B.1.375-like and B.1.258-like cases in 11,977 randomly collected SARS-CoV-2-positive specimens.^6, 8^

The accuracy of the RT-qPCR genotyping was verified against either Sanger sequencing of the receptor-binding domain (RBD) of SARS-CoV-2 surface glycoprotein (S) gene, or by viral whole-genome sequencing on a Nanopore GridION sequencing device. From 236 random samples (27 B.1.1.7-like, 186 B.1.351/P.1-like, and 23 “other” variants), the PCR genotyping results for B.1.1.7-like, B.1.351/P.1-like, and ‘other’ variants were in 88.8% (23 out of 27), 99.5% (185 out of 186), and 100% (23 out of 23) agreement with the SARS-CoV-2 lineages assigned by sequencing.^6, 8^

Within the “other” variant category, Sanger sequencing and/or Illumina sequencing of the RBD of SARS-CoV-2 spike gene on 457 random samples confirmed that 433 (94.7%) were B.1.617.2 cases, 8 (1.8%) were B.1.617.1 cases, 3 (0.7%) were B.1 cases, 1 (0.2%) was a B.1.351/P.1 case, 1 (0.2%) was a P.1 case, and 1 (0.2%) was a B.1.617.3 case, with 10 (1.1%) samples failing lineage assignment.^6, 8^ Accordingly, a Delta case was proxied as any “other” case identified through the RT-qPCR based variant screening.

Within the “other” variant category, Sanger sequencing and/or Illumina sequencing of the RBD of SARS-CoV-2 spike gene on 450 random samples confirmed that 433 (96.2%) were B.1.617.2 cases, 8 (1.8%) were B.1.617.1 cases, 3 (0.7%) were B.1 cases, 1 (0.2%) was a P.1 case, and 1 (0.2%) was a B.1.617.3 case, with 5 (1.1%) samples failing sequencing.^6, 8^ Accordingly, a Delta case was proxied as any “other” case identified through the RT-qPCR based variant screening. All the variant RT-qPCR screening was conducted at the Sidra Medicine Laboratory following standardized protocols.

### Supplementary Section 4. Additional sensitivity analyses

Additional sensitivity analyses were conducted to investigate whether the generated real-world effectiveness estimates could have been biased by an unknown factor. These analyses included:

1. Sensitivity analysis in which study inclusion and exclusion criteria were modified so as to additionally exclude any case or control with a prior infection, that is any person with a PCR-positive test prior to January 1, 2021, the first day of the study (Supplementary Table 5);
2. Sensitivity analysis in which cases and controls were additionally matched by the status of prior infection before study onset, January 1, 2021 (no prior infection, infection in prior 90 days, infection >90 days ago^10, 11^; Supplementary Table 6);
3. Sensitivity analysis in which study inclusion and exclusion criteria were modified so as to additionally include as controls persons who had a PCR-negative test during the study, in addition to the PCR-positive test during the study (Supplementary Table 7). That is, persons with both PCR-positive and PCR-negative tests during the study were included both as cases and as controls, but at different time points;
4. Sensitivity analysis in which study inclusion and exclusion criteria were modified so as to include all PCR-positive and PCR-negative tests for each person, and regardless of the number of PCR-positive or PCR-negative tests each person had during the study (Supplementary Table 8);
5. Sensitivity analysis in which study inclusion and exclusion criteria were modified so as to include all PCR-positive and PCR-negative tests for each person, but all PCR-negative tests for persons included as cases were excluded from analysis. That is, no person was included as both a case and a control (Supplementary Table 9);
6. Sensitivity analysis in which study inclusion and exclusion criteria were modified so as to include all persons vaccinated with a vaccine other than BNT162b2, provided that the PCR test was conducted before receiving the first dose of this vaccine and during the study duration (Supplementary Table 10).

---

**Supplementary Table 1.**
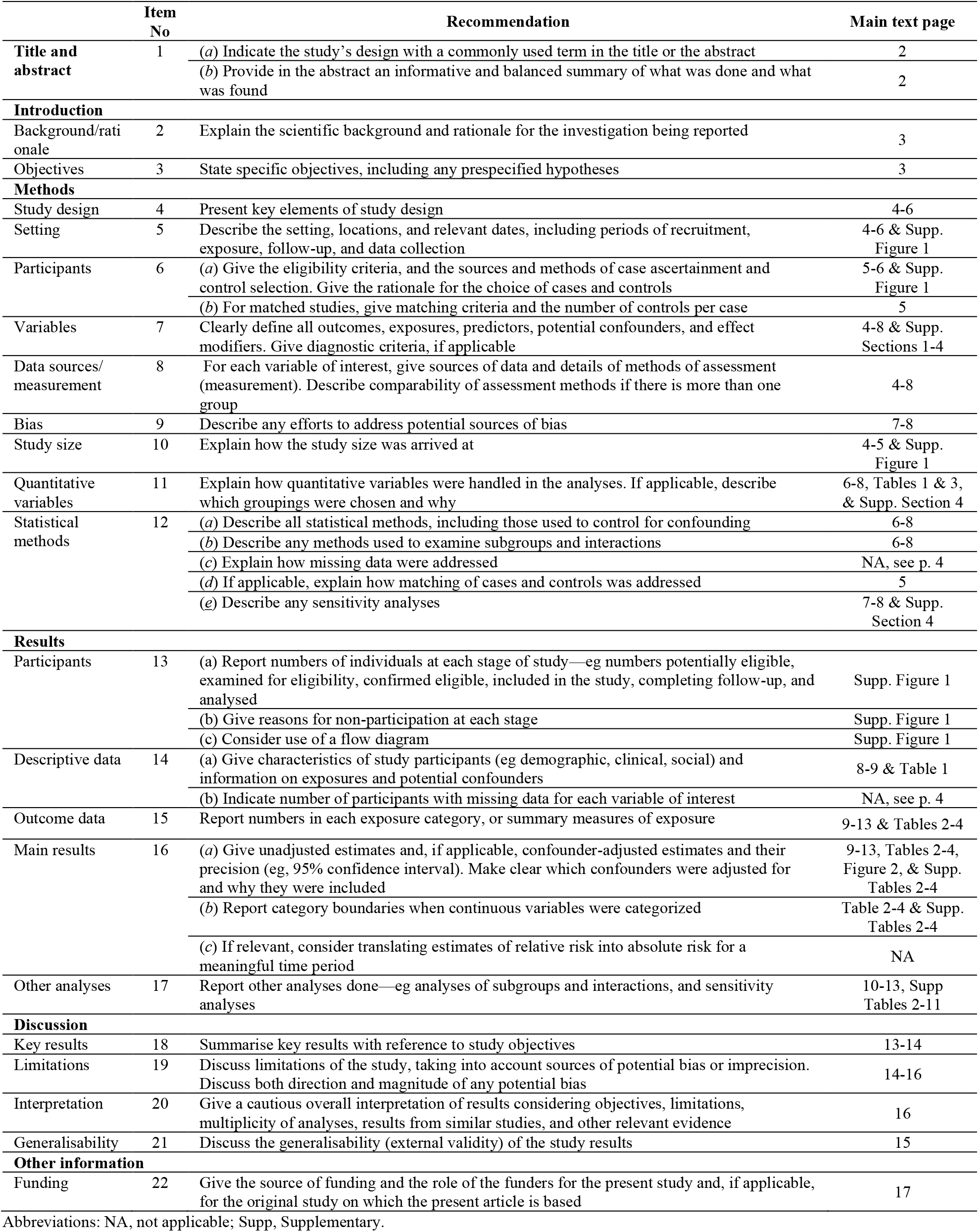
STROBE checklist for case-control studies.

| Supplementary Table 1: Checklist for case-control studies |  |  |  |
| --- | --- | --- | --- |
|  | Item No | Recommendation | Main text page |
| Title and abstract | 1 | (a) Indicate the study's design with a commonly used term in the title or the abstract | 2 |
|  |  | (b) Provide in the abstract an informative and balanced summary of what was done and what was found | 2 |
| Introduction |  |  |  |
| Background/rationale | 2 | Explain the scientific background and rationale for the investigation being reported | 3 |
| Objectives | 3 | State specific objectives, including any prespecified hypotheses | 3 |
| Methods |  |  |  |
| Study design | 4 | Present key elements of study design | 4-6 |
| Setting | 5 | Describe the setting, locations, and relevant dates, including periods of recruitment, exposure, follow-up, and data collection | 4-6 & Supp. Figure 1 |
| Participants | 6 | (a) Give the eligibility criteria, and the sources and methods of case ascertainment and control selection. Give the rationale for the choice of cases and controls | 5-6 & Supp. Figure 1 |
|  |  | (b) For matched studies, give matching criteria and the number of controls per case | 5 |
| Variables | 7 | Clearly define all outcomes, exposures, predictors, potential confounders, and effect modifiers. Give diagnostic criteria, if applicable | 4-8 & Supp. Sections 1-4 |
| Data sources/measurement | 8 | For each variable of interest, give sources of data and details of methods of assessment (measurement). Describe comparability of assessment methods if there is more than one group | 4-8 |
| Bias | 9 | Describe any efforts to address potential sources of bias | 7-8 |
| Study size | 10 | Explain how the study size was arrived at | 4-5 & Supp. Figure 1 |
| Quantitative variables | 11 | Explain how quantitative variables were handled in the analyses. If applicable, describe which groupings were chosen and why | 6-8, Tables 1 & 3, & Supp. Section 4 |
| Statistical methods | 12 | (a) Describe all statistical methods, including those used to control for confounding | 6-8 |
|  |  | (b) Describe any methods used to examine subgroups and interactions | 6-8 |
|  |  | (c) Explain how missing data were addressed | NA, see p. 4 |
|  |  | (d) If applicable, explain how matching of cases and controls was addressed | 5 |
|  |  | (e) Describe any sensitivity analyses | 7-8 & Supp. Section 4 |
| Results |  |  |  |
| Participants | 13 | (a) Report numbers of individuals at each stage of study—eg numbers potentially eligible, examined for eligibility, confirmed eligible, included in the study, completing follow-up, and analysed | Supp. Figure 1 |
|  |  | (b) Give reasons for non-participation at each stage | Supp. Figure 1 |
|  |  | (c) Consider use of a flow diagram | Supp. Figure 1 |
| Descriptive data | 14 | (a) Give characteristics of study participants (eg demographic, clinical, social) and information on exposures and potential confounders | 8-9 & Table 1 |
|  |  | (b) Indicate number of participants with missing data for each variable of interest | NA, see p. 4 |
| Outcome data | 15 | Report numbers in each exposure category, or summary measures of exposure | 9-13 & Tables 2-4 |
| Main results | 16 | (a) Give unadjusted estimates and, if applicable, confounder-adjusted estimates and their precision (eg, 95% confidence interval). Make clear which confounders were adjusted for and why they were included | 9-13, Tables 2-4, Figure 2, & Supp. Tables 2-4 |
|  |  | (b) Report category boundaries when continuous variables were categorized | Table 2-4 & Supp. Tables 2-4 |
|  |  | (c) If relevant, consider translating estimates of relative risk into absolute risk for a meaningful time period | NA |
| Other analyses | 17 | Report other analyses done—eg analyses of subgroups and interactions, and sensitivity analyses | 10-13, Supp. Tables 2-11 |
| Discussion |  |  |  |
| Key results | 18 | Summarise key results with reference to study objectives | 13-14 |
| Limitations | 19 | Discuss limitations of the study, taking into account sources of potential bias or imprecision. Discuss both direction and magnitude of any potential bias | 14-16 |
| Interpretation | 20 | Give a cautious overall interpretation of results considering objectives, limitations, multiplicity of analyses, results from similar studies, and other relevant evidence | 16 |
| Generalisability | 21 | Discuss the generalisability (external validity) of the study results | 15 |
| Other information |  |  |  |
| Funding | 22 | Give the source of funding and the role of the funders for the present study and, if applicable, for the original study on which the present article is based | 17 |

Abbreviations: NA, not applicable; Supp, Supplementary.

**Supplementary Figure 1.**
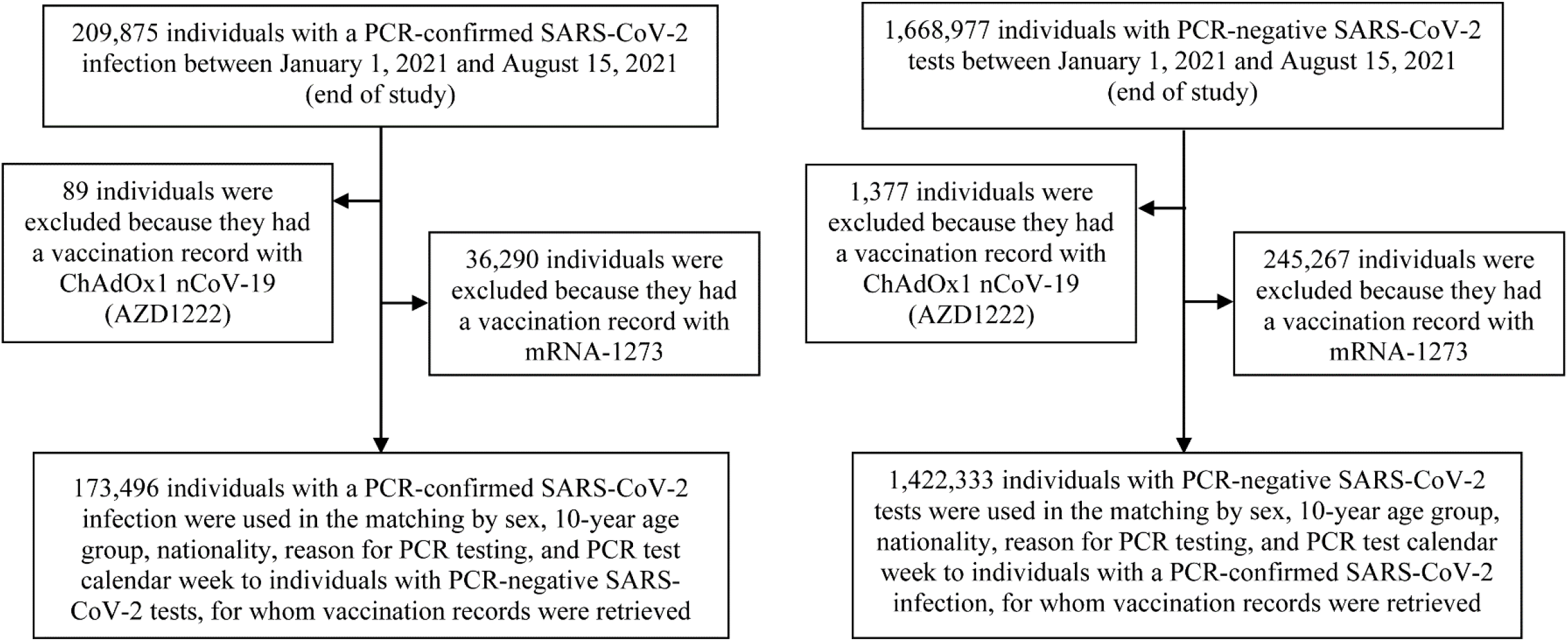
Flowchart describing the population selection process for investigating BNT162b2 vaccine effectiveness.

**Supplementary Table 2.** Effectiveness of the BNT162b2 vaccine against any SARS-CoV-2 infection and against any severe, critical, or fatal COVID-19 disease, stratified by age (<60 years or ≥60 years).

|  | Effectiveness against infection |  |  |  |  | Effectiveness against severity |  |  |  |  |
| --- | --- | --- | --- | --- | --- | --- | --- | --- | --- | --- |
|  | Cases*<br>(PCR-positive) |  | Controls*<br>(PCR-negative) |  | Effectiveness<br>in %<br>(95% CI)<sup>†</sup> | Cases*<br>(Severe, critical, or fatal disease)<sup>‡</sup> |  | Controls*<br>(PCR-negative) |  | Effectiveness<br>in %<br>(95% CI)<sup>†</sup> |
|  | Vaccinated | Unvaccinated | Vaccinated | Unvaccinated |  | Vaccinated | Unvaccinated | Vaccinated | Unvaccinated |  |
| Age <60 years |  |  |  |  |  |  |  |  |  |  |
| 0-13 days after first dose | 3,997 | 137,429 | 3,819 | 137,607 | 0.0<br>(0.0-0.0) | 200 | 3,630 | 212 | 3,618 | 6.0<br>(0.0-23.3) |
| ≥14 days after first dose and no second dose | 2,182 | 137,862 | 3,474 | 136,570 | 37.8<br>(34.3-41.1) | 65 | 3,651 | 235 | 3,481 | 73.6<br>(65.0-80.4) |
| 0-4 weeks after the second dose | 2,925 | 139,313 | 10,139 | 132,099 | 72.6<br>(71.5-73.8) | 18 | 3,681 | 481 | 3,218 | 96.7<br>(94.8-98.1) |
| 5-9 weeks after the second dose | 1,377 | 138,053 | 4,077 | 135,353 | 66.9<br>(64.8-68.9) | 9 | 3,650 | 212 | 3,447 | 96.0<br>(92.2-98.2) |
| 10-14 weeks after the second dose | 833 | 137,664 | 1,909 | 136,588 | 56.7<br>(53.0-60.1) | 3 | 3,624 | 89 | 3,538 | 96.7<br>(90.1-99.3) |
| 15-19 weeks after the second dose | 531 | 137,338 | 734 | 137,135 | 27.8<br>(19.1-35.5) | 4 | 3,626 | 30 | 3,600 | 86.8<br>(62.4-96.6) |
| 20-24 weeks after the second dose | 575 | 137,219 | 478 | 137,316 | 0.0<br>(0.0-0.0) | 1 | 3,623 | 14 | 3,610 | 92.9<br>(53.2-99.8) |
| ≥25 weeks after the second dose | 416 | 137,245 | 369 | 137,292 | 0.0<br>(0.0-2.2) | 1 | 3,631 | 7 | 3,625 | 85.7<br>(0.0-99.7) |
| Age ≥60 years |  |  |  |  |  |  |  |  |  |  |
| 0-13 days after first dose | 197 | 1,471 | 157 | 1,511 | 0.0<br>(0.0-0.0) | 49 | 352 | 39 | 362 | 0.0<br>(0.0-19.1) |
| ≥14 days after first dose and no second dose | 163 | 1,487 | 185 | 1,465 | 13.9<br>(0.0-31.0) | 33 | 366 | 54 | 345 | 42.4<br>(7.0-64.7) |
| 0-4 weeks after the second dose | 216 | 1,621 | 520 | 1,317 | 66.3<br>(59.7-71.8) | 14 | 397 | 117 | 294 | 91.1<br>(84.1-95.4) |
| 5-9 weeks after the second dose | 235 | 1,641 | 533 | 1,343 | 63.9<br>(57.1-69.7) | 12 | 400 | 122 | 290 | 92.9<br>(86.7-96.5) |
| 10-14 weeks after the second dose | 173 | 1,566 | 333 | 1,406 | 53.4<br>(42.9-61.9) | 12 | 376 | 86 | 302 | 88.8<br>(78.9-94.5) |
| 15-19 weeks after the second dose | 50 | 1,490 | 91 | 1,449 | 46.6<br>(23.1-63.2) | 3 | 358 | 21 | 340 | 86.4<br>(53.8-97.4) |
| 20-24 weeks after the second dose | 33 | 1,457 | 45 | 1,445 | 27.3<br>(0.0-55.3) | 0 | 348 | 7 | 341 | 100.0<br>(46.0-100.0) |
| ≥25 weeks after the second dose | 67 | 1,457 | 56 | 1,468 | 0.0<br>(0.0-17.4) | 3 | 348 | 7 | 344 | 57.6<br>(0.0-93.0) |

Abbreviations: CI, confidence interval; PCR, polymerase chain reaction.
\*Cases and controls were matched one-to-one by sex, 10-year age group, nationality, reason for PCR testing, and calendar week of PCR test.
<sup>†</sup>Vaccine effectiveness was estimated using the test-negative, case-control study design.<sup>12,13</sup>
<sup>‡</sup>Severity,<sup>1</sup> criticality,<sup>1</sup> and fatality<sup>2</sup> were defined as per World Health Organization guidelines.

**Supplementary Table 3.**
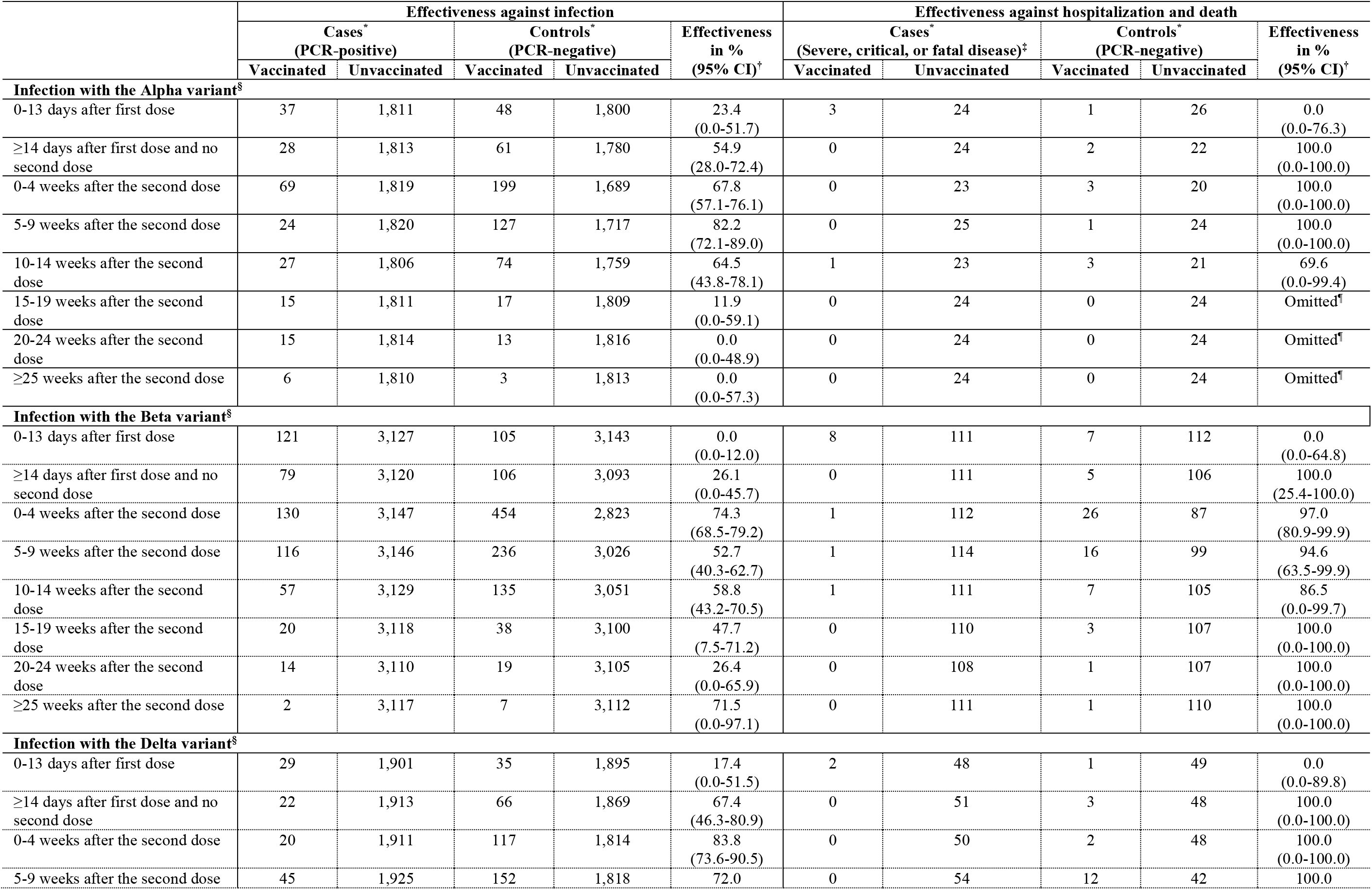
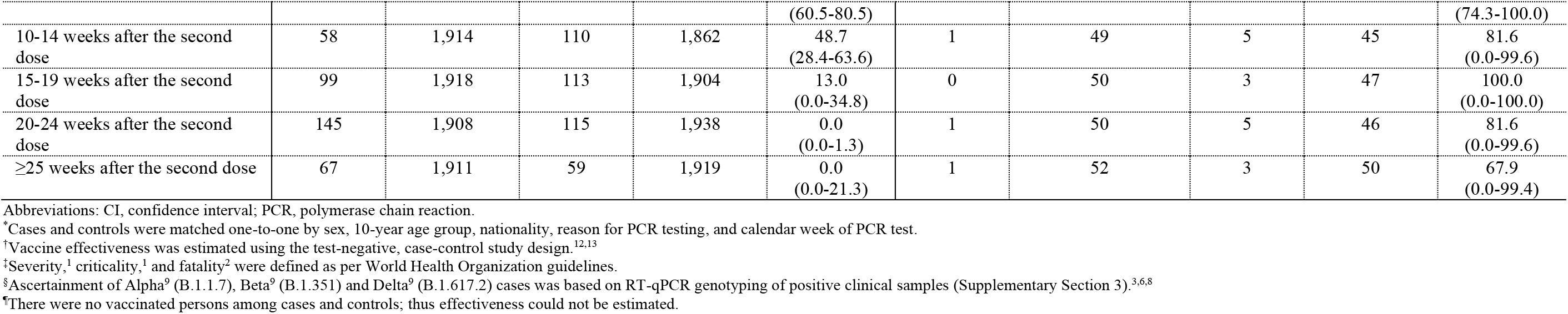
Effectiveness of the BNT162b2 vaccine against each of SARS-CoV-2 Alpha^9^ (B.1.1.7), Beta^9^ (B.1.351) and Delta^9^ (B.1.617.2) variant infections.

**Supplementary Table 4.** Effectiveness of the BNT162b2 vaccine against each of severe COVID-19 disease, critical COVID-19 disease, and fatal COVID-19 disease.

|  | Cases*<br>(COVID-19 disease) |  | Controls*<br>(PCR-negative) |  | Effectiveness in<br>%<br>(95% CI)† |
| --- | --- | --- | --- | --- | --- |
|  | Vaccinated | Unvaccinated | Vaccinated | Unvaccinated |  |
| **Severe disease‡** |  |  |  |  |  |
| 0-13 days after first dose | 221 | 3,363 | 211 | 3,373 | 0.0<br>(0.0-13.9) |
| ≥14 days after first dose and no second dose | 81 | 3,388 | 220 | 3,249 | 64.7<br>(54.0-73.1) |
| 0-4 weeks after the second dose | 30 | 3,437 | 468 | 2,999 | 94.4<br>(91.9-96.3) |
| 5-9 weeks after the second dose | 14 | 3,419 | 267 | 3,166 | 95.1<br>(91.7-97.4) |
| 10-14 weeks after the second dose | 11 | 3,378 | 139 | 3,250 | 92.4<br>(85.9-96.3) |
| 15-19 weeks after the second dose | 6 | 3,364 | 42 | 3,328 | 85.9<br>(66.5-95.1) |
| ≥20 weeks after the second dose | 4 | 3,350 | 22 | 3,332 | 81.9<br>(46.7-95.5) |
| **Critical disease‡** |  |  |  |  |  |
| 0-13 days after first dose | 27 | 609 | 39 | 597 | 32.1<br>(0.0-60.6) |
| ≥14 days after first dose and no second dose | 16 | 619 | 66 | 569 | 77.7<br>(60.5-88.1) |
| 0-4 weeks after the second dose | 2 | 630 | 127 | 505 | 98.7<br>(95.3-99.8) |
| 5-9 weeks after the second dose | 6 | 620 | 65 | 561 | 91.6<br>(80.6-97.1) |
| 10-14 weeks after the second dose | 4 | 612 | 35 | 581 | 89.2<br>(69.3-97.2) |
| 15-19 weeks after the second dose | 1 | 610 | 9 | 602 | 89.0<br>(20.3-99.8) |
| ≥20 weeks after the second dose | 1 | 604 | 3 | 602 | 66.8<br>(0.0-99.4) |
| **Fatal disease‡** |  |  |  |  |  |
| 0-13 days after first dose | 15 | 219 | 10 | 224 | 0.0<br>(0.0-37.1) |
| ≥14 days after first dose and no second dose | 11 | 222 | 30 | 203 | 66.5<br>(28.9-85.2) |
| 0-4 weeks after the second dose | 5 | 230 | 62 | 173 | 93.9<br>(84.5-98.1) |
| 5-9 weeks after the second dose | 5 | 232 | 42 | 195 | 90.0<br>(74.0-97.0) |
| 10-14 weeks after the second dose | 2 | 225 | 27 | 200 | 93.4<br>(73.1-99.2) |
| 15-19 weeks after the second dose | 1 | 217 | 5 | 213 | 80.4<br>(0.0-99.6) |
| ≥20 weeks after the second dose | 0 | 212 | 0 | 212 | Omitted§ |

Abbreviations: CI, confidence interval; PCR, polymerase chain reaction.
\*Cases and controls were matched one-to-one by sex, 10-year age group, nationality, reason for PCR testing, and calendar week of PCR test.
†Vaccine effectiveness was estimated using the test-negative, case-control study design.<sup>12,13</sup>
‡Severity,<sup>1</sup> criticality,<sup>1</sup> and fatality<sup>2</sup> were defined as per World Health Organization guidelines.
§There were no vaccinated persons among cases and controls; thus effectiveness could not be estimated.

**Supplementary Table 5.** Sensitivity analysis in which study inclusion and exclusion criteria were modified so as to additionally exclude any case or control with a prior infection, that is any person with a PCR-positive test prior to January 1, 2021, the first day of the study. Effectiveness of the BNT162b2 vaccine against any SARS-CoV-2 infection and against any severe, critical, or fatal COVID-19 disease.

|  | Effectiveness against infection |  |  |  |  | Effectiveness against hospitalization and death |  |  |  |  |
| --- | --- | --- | --- | --- | --- | --- | --- | --- | --- | --- |
|  | Cases*<br>(PCR-positive) |  | Controls*<br>(PCR-negative) |  | Effectiveness<br>in %<br>(95% CI)<sup>†</sup> | Cases*<br>(Severe, critical, or fatal disease)* |  | Controls*<br>(PCR-negative) |  | Effectiveness<br>in %<br>(95% CI)<sup>†</sup> |
|  | Vaccinated | Unvaccinated | Vaccinated | Unvaccinated |  | Vaccinated | Unvaccinated | Vaccinated | Unvaccinated |  |
| 0-13 days after first dose | 4,055 | 136,097 | 3,829 | 136,323 | 0.0<br>(0.0-0.0) | 246 | 3,943 | 264 | 3,925 | 7.2<br>(0.0-22.8) |
| ≥14 days after first dose and no second dose | 2,274 | 136,527 | 3,411 | 135,390 | 33.9<br>(30.2-37.4) | 101 | 3,987 | 292 | 3,796 | 67.1<br>(58.4-74.1) |
| 0-4 weeks after the second dose | 3,078 | 138,103 | 10,181 | 131,000 | 71.3<br>(70.1-72.5) | 30 | 4,039 | 579 | 3,490 | 95.5<br>(93.5-97.0) |
| 5-9 weeks after the second dose | 1,564 | 136,923 | 4,395 | 134,092 | 65.1<br>(63.0-67.1) | 21 | 4,020 | 336 | 3,705 | 94.2<br>(91.0-96.5) |
| 10-14 weeks after the second dose | 978 | 136,431 | 2,203 | 135,206 | 56.0<br>(52.5-59.2) | 14 | 3,982 | 170 | 3,826 | 92.1<br>(86.3-95.8) |
| 15-19 weeks after the second dose | 566 | 135,982 | 790 | 135,758 | 28.5<br>(20.2-35.9) | 8 | 3,946 | 63 | 3,891 | 87.5<br>(73.7-94.8) |
| 20-24 weeks after the second dose | 591 | 135,894 | 522 | 135,963 | 0.0<br>(0.0-0.0) | 1 | 3,941 | 23 | 3,919 | 95.7<br>(73.3-99.9) |
| ≥25 weeks after the second dose | 472 | 135,864 | 416 | 135,920 | 0.0<br>(0.0-0.7) | 4 | 3,929 | 12 | 3,921 | 66.7<br>(0.0-92.2) |

Abbreviations: CI, confidence interval; PCR, polymerase chain reaction.
\*Cases and controls were matched one-to-one by sex, 10-year age group, nationality, reason for PCR testing, and calendar week of PCR test.
<sup>†</sup>Vaccine effectiveness was estimated using the test-negative, case-control study design.<sup>12,13</sup>
<sup>‡</sup>Severity,<sup>1</sup> criticality,<sup>1</sup> and fatality<sup>2</sup> were defined as per World Health Organization guidelines.

**Supplementary Table 6.** Sensitivity analysis in which the cases and controls were additionally matched by the status of prior infection before study onset, January 1, 2021 (no prior infection, infection in prior 90 days, infection >90 days ago^10, 11^). Effectiveness of the BNT162b2 vaccine against any SARS-CoV-2 infection and against any severe, critical, or fatal COVID-19 disease.

|  | Effectiveness against infection |  |  |  |  | Effectiveness against hospitalization and death |  |  |  |  |
| --- | --- | --- | --- | --- | --- | --- | --- | --- | --- | --- |
|  | Cases*<br>(PCR-positive) |  | Controls*<br>(PCR-negative) |  | Effectiveness<br>in %<br>(95% CI)<sup>†</sup> | Cases*<br>(Severe, critical, or fatal disease)* |  | Controls*<br>(PCR-negative) |  | Effectiveness<br>in %<br>(95% CI)<sup>†</sup> |
|  | Vaccinated | Unvaccinated | Vaccinated | Unvaccinated |  | Vaccinated | Unvaccinated | Vaccinated | Unvaccinated |  |
| 0-13 days after first dose | 4,090 | 137,646 | 3,907 | 137,829 | 0.0<br>(0.0-0.0) | 238 | 3,979 | 264 | 3,953 | 10.4<br>(0.0-25.5) |
| ≥14 days after first dose and no second dose | 2,305 | 138,004 | 3,510 | 136,799 | 34.9<br>(31.3-38.3) | 97 | 4,002 | 332 | 3,767 | 72.5<br>(65.2-78.4) |
| 0-4 weeks after the second dose | 3,099 | 139,608 | 10,355 | 132,352 | 71.6<br>(70.4-72.8) | 31 | 4,037 | 602 | 3,466 | 95.6<br>(93.6-97.0) |
| 5-9 weeks after the second dose | 1,594 | 138,417 | 4,423 | 135,588 | 64.7<br>(62.6-66.7) | 21 | 4,034 | 324 | 3,731 | 94.0<br>(90.7-96.3) |
| 10-14 weeks after the second dose | 999 | 137,935 | 2,162 | 136,772 | 54.2<br>(50.6-57.5) | 14 | 3,985 | 179 | 3,820 | 92.5<br>(87.1-96.0) |
| 15-19 weeks after the second dose | 578 | 137,527 | 794 | 137,311 | 27.3<br>(19.0-34.8) | 6 | 3,960 | 54 | 3,912 | 89.0<br>(74.5-96.1) |
| 20-24 weeks after the second dose | 600 | 137,443 | 530 | 137,513 | 0.0<br>(0.0-0.0) | 1 | 3,949 | 19 | 3,931 | 94.8<br>(67.0-99.9) |
| ≥25 weeks after the second dose | 477 | 137,394 | 405 | 137,466 | 0.0<br>(0.0-0.7) | 4 | 3,944 | 12 | 3,936 | 66.7<br>(0.0-92.2) |

Abbreviations: CI, confidence interval; PCR, polymerase chain reaction.
\*Cases and controls were matched one-to-one by sex, 10-year age group, nationality, reason for PCR testing, calendar week of PCR test, and status of prior infection.
<sup>†</sup>Vaccine effectiveness was estimated using the test-negative, case-control study design.<sup>12,13</sup>
<sup>‡</sup>Severity,<sup>1</sup> criticality,<sup>1</sup> and fatality<sup>2</sup> were defined as per World Health Organization guidelines.

**Supplementary Table 7.** Sensitivity analysis in which study inclusion and exclusion criteria were modified so as to additionally include as controls persons who had a PCR-negative test during the study, in addition to their PCR positive test during the study. That is, persons with both PCR-positive and PCR-negative tests during the study, January 1, 2021 to August 15, 2021, were included both as cases and as controls, but at different time points. Effectiveness of the BNT162b2 vaccine against any SARS-CoV-2 infection and against any severe, critical, or fatal COVID-19 disease.

|  | Effectiveness against infection |  |  |  |  | Effectiveness against hospitalization and death |  |  |  |  |
| --- | --- | --- | --- | --- | --- | --- | --- | --- | --- | --- |
|  | Cases*<br>(PCR-positive) |  | Controls*<br>(PCR-negative) |  | Effectiveness<br>in %<br>(95% CI)<sup>†</sup> | Cases*<br>(Severe, critical, or fatal disease)<sup>‡</sup> |  | Controls*<br>(PCR-negative) |  | Effectiveness<br>in %<br>(95% CI)<sup>†</sup> |
|  | Vaccinated | Unvaccinated | Vaccinated | Unvaccinated |  | Vaccinated | Unvaccinated | Vaccinated | Unvaccinated |  |
| 0-13 days after first dose | 4,441 | 143,720 | 3,856 | 144,305 | 0.0<br>(0.0-0.0) | 265 | 4,112 | 264 | 4,113 | 0.0<br>(0.0-16.1) |
| ≥14 days after first dose and no second dose | 2,564 | 144,171 | 3,845 | 142,890 | 33.9<br>(30.5-37.2) | 108 | 4,145 | 296 | 3,957 | 65.2<br>(56.2-72.4) |
| 0-4 weeks after the second dose | 3,277 | 145,159 | 10,059 | 138,377 | 68.9<br>(67.7-70.2) | 33 | 4,170 | 508 | 3,695 | 94.2<br>(91.8-96.1) |
| 5-9 weeks after the second dose | 1,682 | 144,272 | 4,315 | 141,639 | 61.7<br>(59.5-63.9) | 23 | 4,154 | 281 | 3,896 | 92.3<br>(88.2-95.2) |
| 10-14 weeks after the second dose | 1,056 | 143,991 | 2,111 | 142,936 | 50.3<br>(46.5-53.9) | 16 | 4,144 | 178 | 3,982 | 91.4<br>(85.5-95.2) |
| 15-19 weeks after the second dose | 611 | 143,707 | 809 | 143,509 | 24.6<br>(16.1-32.2) | 7 | 4,115 | 42 | 4,080 | 83.5<br>(62.8-93.7) |
| 20-24 weeks after the second dose | 616 | 143,620 | 545 | 143,691 | 0.0<br>(0.0-0.0) | 1 | 4,114 | 20 | 4,095 | 95.0<br>(68.8-99.9) |
| ≥25 weeks after the second dose | 499 | 143,602 | 404 | 143,697 | 0.0<br>(0.0-0.0) | 4 | 4,112 | 12 | 4,104 | 66.7<br>(0.0-92.2) |

Abbreviations: CI, confidence interval; PCR, polymerase chain reaction.
\*Cases and controls were matched one-to-one by sex, 10-year age group, nationality, reason for PCR testing, and calendar week of PCR test.
<sup>†</sup>Vaccine effectiveness was estimated using the test-negative, case-control study design.<sup>12,13</sup>
<sup>‡</sup>Severity,<sup>1</sup> criticality,<sup>1</sup> and fatality<sup>2</sup> were defined as per World Health Organization guidelines.

**Supplementary Table 8.** Sensitivity analysis in which study inclusion and exclusion criteria were modified so as to include all PCR-positive and PCR-negative tests for each person, and regardless of the number of PCR-positive or PCR-negative tests each person had during the study, January 1, 2021 to August 15, 2021. Effectiveness of the BNT162b2 vaccine against any SARS-CoV-2 infection and against any severe, critical, or fatal COVID-19 disease.

|  | Effectiveness against infection |  |  |  |  | Effectiveness against hospitalization and death |  |  |  |  |
| --- | --- | --- | --- | --- | --- | --- | --- | --- | --- | --- |
|  | Cases*<br>(PCR-positive) |  | Controls*<br>(PCR-negative) |  | Effectiveness<br>in %<br>(95% CI)<sup>†</sup> | Cases*<br>(Severe, critical, or fatal disease)* |  | Controls*<br>(PCR-negative) |  | Effectiveness<br>in %<br>(95% CI)<sup>†</sup> |
|  | Vaccinated | Unvaccinated | Vaccinated | Unvaccinated |  | Vaccinated | Unvaccinated | Vaccinated | Unvaccinated |  |
| 0-13 days after first dose | 4,981 | 173,946 | 5,513 | 173,414 | 9.9<br>(6.3-13.4) | 298 | 5,239 | 331 | 5,206 | 10.5<br>(0.0-24.1) |
| ≥14 days after first dose and no second dose | 4,070 | 174,240 | 5,920 | 172,390 | 32.0<br>(29.2-34.7) | 183 | 5,253 | 424 | 5,012 | 58.8<br>(50.6-65.7) |
| 0-4 weeks after the second dose | 3,988 | 175,262 | 14,301 | 164,949 | 73.8<br>(72.8-74.7) | 42 | 5,308 | 754 | 4,596 | 95.2<br>(93.4-96.6) |
| 5-9 weeks after the second dose | 2,143 | 174,426 | 7,865 | 168,704 | 73.6<br>(72.3-74.9) | 32 | 5,297 | 460 | 4,869 | 93.6<br>(90.8-95.7) |
| 10-14 weeks after the second dose | 1,393 | 174,105 | 4,569 | 170,929 | 70.1<br>(68.2-71.8) | 30 | 5,283 | 304 | 5,009 | 90.6<br>(86.3-93.8) |
| 15-19 weeks after the second dose | 863 | 173,751 | 1,900 | 172,714 | 54.9<br>(51.0-58.4) | 13 | 5,245 | 92 | 5,166 | 86.1<br>(75.0-92.9) |
| 20-24 weeks after the second dose | 840 | 173,653 | 1,200 | 173,293 | 30.1<br>(23.6-36.1) | 3 | 5,244 | 38 | 5,209 | 92.2<br>(75.3-98.5) |
| ≥25 weeks after the second dose | 687 | 173,655 | 887 | 173,455 | 22.6<br>(14.4-30.1) | 7 | 5,238 | 25 | 5,220 | 72.1<br>(33.6-89.8) |

Abbreviations: CI, confidence interval; PCR, polymerase chain reaction.
\*Cases and controls were matched one-to-one by sex, 10-year age group, nationality, reason for PCR testing, and calendar week of PCR test.
<sup>†</sup>Vaccine effectiveness was estimated using the test-negative, case-control study design.<sup>12,13</sup>
<sup>‡</sup>Severity,<sup>1</sup> criticality,<sup>1</sup> and fatality<sup>2</sup> were defined as per World Health Organization guidelines.

**Supplementary Table 9.**
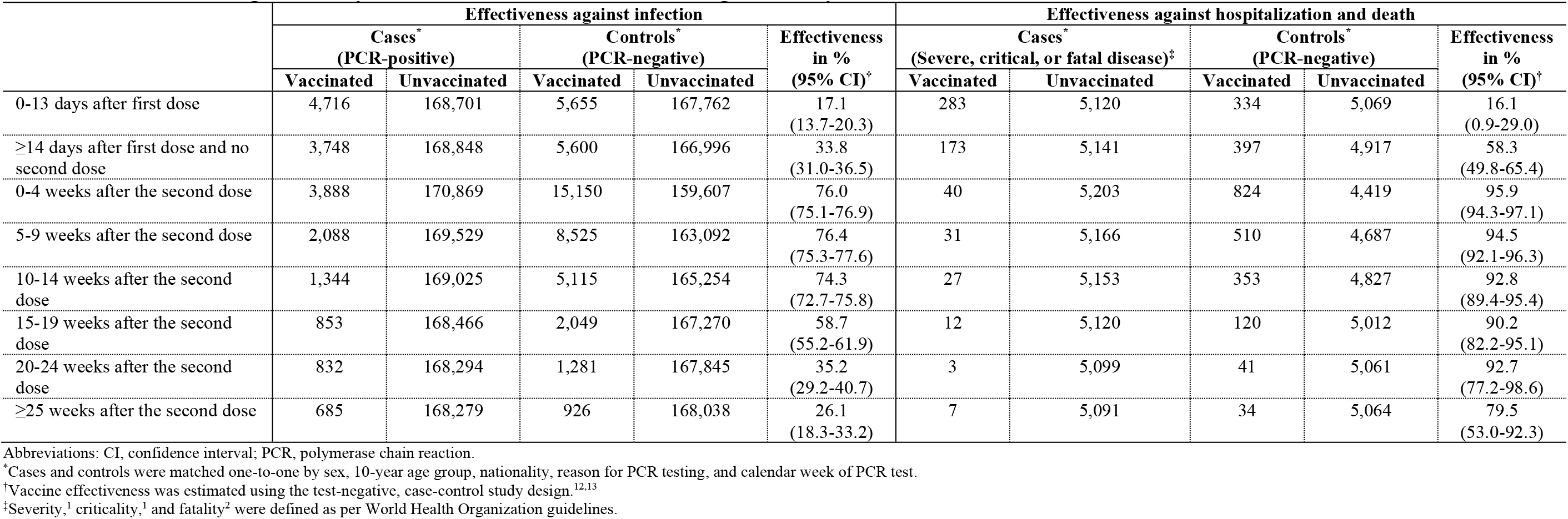
Sensitivity analysis in which study inclusion and exclusion criteria were modified so as to include all PCR-positive and PCR-negative tests for each person, and regardless of the number of PCR-positive or PCR-negative tests each person had during the study, January 1, 2021 to August 15, 2021. However, all PCR-negative tests for persons included as cases were excluded from analysis. That is, no person was included as both a case and a control. Effectiveness of the BNT162b2 vaccine against any SARS-CoV-2 infection and against any severe, critical, or fatal COVID-19 disease.

**Supplementary Table 10.** Sensitivity analysis in which study inclusion and exclusion criteria were modified so as to additionally include all persons vaccinated with a vaccine other than BNT162b2 provided that the PCR test was conducted before receiving the first dose of this vaccine, and during the study, January 1, 2021 to August 15, 2021. Effectiveness of the BNT162b2 vaccine against any SARS-CoV-2 infection and against any severe, critical, or fatal COVID-19 disease.

|  | Effectiveness against infection |  |  |  |  | Effectiveness against hospitalization and death |  |  |  |  |
| --- | --- | --- | --- | --- | --- | --- | --- | --- | --- | --- |
|  | Cases*<br>(PCR-positive) |  | Controls*<br>(PCR-negative) |  | Effectiveness<br>in %<br>(95% CI)<sup>†</sup> | Cases*<br>(Severe, critical, or fatal disease)* |  | Controls*<br>(PCR-negative) |  | Effectiveness<br>in %<br>(95% CI)<sup>†</sup> |
|  | Vaccinated | Unvaccinated | Vaccinated | Unvaccinated |  | Vaccinated | Unvaccinated | Vaccinated | Unvaccinated |  |
| 0-13 days after first dose | 4,302 | 166,610 | 4,038 | 166,874 | 0.0<br>(0.0-0.0) | 257 | 5,276 | 248 | 5,285 | 0.0<br>(0.0-13.5) |
| ≥14 days after first dose and no second dose | 2,372 | 167,125 | 3,747 | 165,750 | 37.2<br>(33.9-40.4) | 105 | 5,320 | 295 | 5,130 | 65.7<br>(56.8-72.9) |
| 0-4 weeks after the second dose | 3,137 | 168,828 | 11,295 | 160,670 | 73.6<br>(72.5-74.6) | 34 | 5,364 | 662 | 4,736 | 95.5<br>(93.6-96.9) |
| 5-9 weeks after the second dose | 1,613 | 167,457 | 4,803 | 164,267 | 67.1<br>(65.1-68.9) | 20 | 5,350 | 353 | 5,017 | 94.7<br>(91.7-96.8) |
| 10-14 weeks after the second dose | 1,024 | 167,061 | 2,275 | 165,810 | 55.3<br>(51.9-58.5) | 16 | 5,307 | 189 | 5,134 | 91.8<br>(86.3-95.4) |
| 15-19 weeks after the second dose | 589 | 166,610 | 858 | 166,341 | 31.5<br>(23.8-38.4) | 7 | 5,277 | 46 | 5,238 | 84.9<br>(66.3-94.3) |
| 20-24 weeks after the second dose | 607 | 166,536 | 558 | 166,585 | 0.0<br>(0.0-3.2) | 1 | 5,266 | 20 | 5,247 | 95.1<br>(68.8-99.9) |
| ≥25 weeks after the second dose | 489 | 166,532 | 399 | 166,622 | 0.0<br>(0.0-0.0) | 4 | 5,271 | 11 | 5,264 | 63.7<br>(0.0-91.6) |

Abbreviations: CI, confidence interval; PCR, polymerase chain reaction.
\*Cases and controls were matched one-to-one by sex, 10-year age group, nationality, reason for PCR testing, and calendar week of PCR test.
<sup>†</sup>Vaccine effectiveness was estimated using the test-negative, case-control study design.<sup>12,13</sup>
<sup>‡</sup>Severity,<sup>1</sup> criticality,<sup>1</sup> and fatality<sup>2</sup> were defined as per World Health Organization guidelines.

**Supplementary Table 11.** Effectiveness of the BNT162b2 vaccine against any SARS-CoV-2 infection, symptomatic SARS-CoV-2 infection, or asymptomatic SARS-CoV-2 infection, with effectiveness estimated using multivariable logistic regression analysis of associations with a PCR-positive test, January 1, 2021 to August 15, 2021, adjusting for sex, age, nationality, reason for PCR testing, prior infection, and calendar week of PCR test*.

|  | Original sample size | SARS-CoV-2 positive |  | Univariable regression analysis |  | Multivariable regression analysis |  | Vaccine effectiveness<sup>†</sup> |
| --- | --- | --- | --- | --- | --- | --- | --- | --- |
|  | N (%) | N (%) | p-value | OR (95% CI) | p-value | AOR (95% CI) | p-value | % (95% CI) |
| **Any SARS-CoV-2 infection** |  |  |  |  |  |  |  |  |
| Unvaccinated | 1,318,985 (82.7) | 156,427 (11.9) | <0.001 | 1.00 |  | 1.00 |  |  |
| <14 days after dose 1 and no dose 2 | 22,957 (1.4) | 5,216 (22.7) |  | 2.19 (2.12-2.25) | <0.001 | 1.10 (1.07-1.14) | <0.001 | 0.0 (0.0-0.0) |
| ≥14 days after dose 1 and no dose 2 | 18,196 (1.1) | 2,990 (16.4) |  | 1.46 (1.40-1.52) | <0.001 | 0.68 (0.65-0.71) | <0.001 | 31.8 (28.8-34.7) |
| 0-4 weeks after dose 2 | 87,656 (5.5) | 3,607 (4.1) |  | 0.32 (0.31-0.33) | <0.001 | 0.23 (0.22-0.24) | <0.001 | 77.2 (76.4-78.0) |
| 5-9 weeks after dose 2 | 56,353 (3.5) | 1,957 (3.5) |  | 0.27 (0.26-0.28) | <0.001 | 0.26 (0.25-0.27) | <0.001 | 73.9 (72.6-75.1) |
| 10-14 weeks after dose 2 | 40,119 (2.5) | 1,253 (3.1) |  | 0.24 (0.23-0.25) | <0.001 | 0.33 (0.31-0.35) | <0.001 | 67.2 (65.2-69.2) |
| 15-19 weeks after dose 2 | 26,425 (1.7) | 710 (2.7) |  | 0.21 (0.19-0.22) | <0.001 | 0.48 (0.45-0.52) | <0.001 | 51.6 (47.7-55.3) |
| 20-24 weeks after dose 2 | 16,594 (1.0) | 726 (4.4) |  | 0.34 (0.32-0.37) | <0.001 | 0.94 (0.87-1.01) | 0.108 | 6.3 (0.0-13.5) |
| ≥25 weeks after dose 2 | 8,544 (0.5) | 610 (7.1) |  | 0.57 (0.53-0.62) | <0.001 | 1.65 (1.51-1.80) | <0.001 | 0.0 (0.0-0.0) |
| **Symptomatic SARS-CoV-2 infection<sup>‡</sup>** |  |  |  |  |  |  |  |  |
| Unvaccinated | 137,557 (81.7) | 46,566 (33.9) | <0.001 | 1.00 |  | 1.00 |  |  |
| <14 days after dose 1 and no dose 2 | 5,912 (3.5) | 2,617 (44.3) |  | 1.55 (1.47-1.64) | <0.001 | 1.05 (0.99-1.11) | 0.128 | 0.0 (0.0-1.3) |
| ≥14 days after dose 1 and no dose 2 | 4,946 (2.9) | 1,456 (29.4) |  | 0.82 (0.77-0.87) | <0.001 | 0.52 (0.48-0.55) | <0.001 | 48.5 (44.9-51.8) |
| 0-4 weeks after dose 2 | 7,311 (4.3) | 1,025 (14.0) |  | 0.32 (0.30-0.34) | <0.001 | 0.18 (0.17-0.19) | <0.001 | 82.1 (80.7-83.3) |
| 5-9 weeks after dose 2 | 4,796 (2.8) | 745 (15.5) |  | 0.36 (0.33-0.39) | <0.001 | 0.26 (0.24-0.28) | <0.001 | 73.9 (71.6-76.0) |
| 10-14 weeks after dose 2 | 3,505 (2.1) | 481 (13.7) |  | 0.31 (0.28-0.34) | <0.001 | 0.36 (0.33-0.40) | <0.001 | 63.8 (59.7-67.4) |
| 15-19 weeks after dose 2 | 1,974 (1.2) | 231 (11.7) |  | 0.26 (0.23-0.30) | <0.001 | 0.60 (0.52-0.70) | <0.001 | 39.6 (30.0-47.9) |
| 20-24 weeks after dose 2 | 1,408 (0.8) | 219 (15.6) |  | 0.36 (0.31-0.42) | <0.001 | 1.17 (1.00-1.36) | 0.046 | 0.0 (0.0-0.0) |
| ≥25 weeks after dose 2 | 909 (0.5) | 215 (23.7) |  | 0.61 (0.52-0.71) | <0.001 | 2.42 (2.05-2.85) | <0.001 | 0.0 (0.0-0.0) |
| **Asymptomatic SARS-CoV-2 infection<sup>§</sup>** |  |  |  |  |  |  |  |  |
| Unvaccinated | 901,253 (82.7) | 62,436 (6.9) | <0.001 | 1.00 |  | 1.00 |  |  |
| <14 days after dose 1 and no dose 2 | 10,304 (0.9) | 1,171 (11.4) |  | 1.72 (1.62-1.83) | <0.001 | 1.02 (0.96-1.09) | 0.487 | 0.0 (0.0-4.1) |
| ≥14 days after dose 1 and no dose 2 | 8,368 (0.8) | 688 (8.2) |  | 1.20 (1.11-1.30) | <0.001 | 0.85 (0.78-0.92) | <0.001 | 15.2 (8.0-21.8) |
| 0-4 weeks after dose 2 | 56,707 (5.2) | 1,288 (2.3) |  | 0.31 (0.30-0.33) | <0.001 | 0.30 (0.29-0.32) | <0.001 | 69.7 (67.9-71.4) |
| 5-9 weeks after dose 2 | 40,613 (3.7) | 735 (1.8) |  | 0.25 (0.23-0.27) | <0.001 | 0.31 (0.29-0.33) | <0.001 | 69.0 (66.6-71.3) |
| 10-14 weeks after dose 2 | 31,078 (2.9) | 534 (1.7) |  | 0.23 (0.22-0.26) | <0.001 | 0.38 (0.35-0.42) | <0.001 | 61.8 (58.3-65.0) |
| 15-19 weeks after dose 2 | 21,635 (2.0) | 358 (1.7) |  | 0.23 (0.20-0.25) | <0.001 | 0.56 (0.50-0.62) | <0.001 | 44.3 (38.0-50.0) |
| 20-24 weeks after dose 2 | 13,403 (1.2) | 384 (2.9) |  | 0.40 (0.36-0.44) | <0.001 | 1.06 (0.96-1.18) | 0.253 | 0.0 (0.0-4.3) |
| ≥25 weeks after dose 2 | 6,641 (0.6) | 293 (4.4) |  | 0.62 (0.55-0.70) | <0.001 | 1.69 (1.49-1.91) | <0.001 | 0.0 (0.0-0.0) |

Abbreviations: AOR: adjusted odds ratio; CI, confidence interval; OR: odds ratio.
<sup>‡</sup>Analyses were conducted on the full sample including 171,352 individuals with a first PCR-positive test and 1,363,261 individuals with a first PCR-negative test. Variables were included in categorical form as follows: sex (male, female), age (0-9, 10-19, 20-29, 30-39, 40-49, 50-59, 60-69, and 70+ years), nationality (Bangladeshi, Egyptians, Filipinos, Indians, Nepalese, Pakistani, Qataris, Sri Lankans, Sudanese, and other nationalities), reason for PCR testing (clinical suspicion, contact tracing, healthcare routine testing, survey, port of entry, pre-travel, individual request, and other), calendar week of PCR test starting from January 1, 2021, and prior infection (no prior infection, infection in prior 90 days, infection >90 days ago<sup>10,11</sup>).
<sup>†</sup>Vaccine effectiveness was calculated using the equation: 1 – AOR, that is assuming odds ratio approximates risk ratio for rare outcomes.
<sup>‡</sup>A symptomatic infection was defined as a PCR-positive test conducted because of clinical suspicion due to presence of symptoms compatible with a respiratory tract infection.
<sup>§</sup>An asymptomatic infection was defined as a PCR-positive test conducted with no reported presence of symptoms compatible with a respiratory tract infection. That is, the PCR testing was done as part of a survey, for pre-travel requirement, or at port of entry into the country.

## Notes

### Competing Interest Statement

The authors have declared no competing interest.

### Author Declarations

The study was approved by the Hamad Medical Corporation and Weill Cornell Medicine-Qatar Institutional Review Boards with waiver of informed consent.

## References

1. Polack FP, Thomas SJ, Kitchin N, et al. Safety and Efficacy of the BNT162b2 mRNA Covid-19 Vaccine. N Engl J Med 2020.

2. Abu-Raddad LJ, Chemaitelly H, Butt AA, National Study Group for Covid-19 Vaccination. Effectiveness of the BNT162b2 Covid-19 Vaccine against the B.1.1.7 and B.1.351 Variants. N Engl J Med 2021.

3. Baden LR, El Sahly HM, Essink B, et al. Efficacy and Safety of the mRNA-1273 SARS-CoV-2 Vaccine. N Engl J Med 2021;384:403–16.

4. Chemaitelly H, Yassine HM, Benslimane FM, et al. mRNA-1273 COVID-19 vaccine effectiveness against the B.1.1.7 and B.1.351 variants and severe COVID-19 disease in Qatar. Nat Med 2021.

5. Ministry of Public Health. National Covid-19 Testing and Vaccination Program Data. https://covid19.moph.gov.qa/EN/Pages/Vaccination-Program-Data.aspx. Access date August 19, 2021. 2021.

6. Coronavirus (COVID-19) vaccinations. Our World in Data. 2021. (Accessed August 20, 2021, at https://ourworldindata.org/covid-vaccinations.)

7. World Health Organization. Tracking SARS-CoV-2 variants. Available from: https://www.who.int/en/activities/tracking-SARS-CoV-2-variants/. Accessed on: June 5, 2021. 2021.

8. Qatar viral genome sequencing data. Data on randomly collected samples. https://www.gisaid.org/phylodynamics/global/nextstrain/. 2021. at https://www.gisaid.org/phylodynamics/global/nextstrain/.)

9. Benslimane FM, Al Khatib HA, Al-Jamal O, et al. One year of SARS-CoV-2: Genomic characterization of COVID-19 outbreak in Qatar. medRxiv 2021:2021.05.19.21257433.

10. Hasan MR, Kalikiri MKR, Mirza F, et al. Real-Time SARS-CoV-2 Genotyping by High-Throughput Multiplex PCR Reveals the Epidemiology of the Variants of Concern in Qatar. medRxiv 2021:2021.07.18.21260718.

11. Abu-Raddad LJ, Chemaitelly H, Ayoub HH, et al. Characterizing the Qatar advanced-phase SARS-CoV-2 epidemic. Scientific Reports 2021;11:6233.

12. Planning and Statistics Authority-State of Qatar. Qatar Monthly Statistics. Available from: https://www.psa.gov.qa/en/pages/default.aspx. Accessed on: May 26, 2020. 2020.

13. Bertollini R, Chemaitelly H, Yassine HM, Al-Thani MH, Al-Khal A, Abu-Raddad LJ. Associations of Vaccination and of Prior Infection With Positive PCR Test Results for SARS-CoV-2 in Airline Passengers Arriving in Qatar. JAMA 2021.

14. Jackson ML, Nelson JC. The test-negative design for estimating influenza vaccine effectiveness. Vaccine 2013;31:2165–8.

15. Verani JR, Baqui AH, Broome CV, et al. Case-control vaccine effectiveness studies: Preparation, design, and enrollment of cases and controls. Vaccine 2017;35:3295–302.

16. Lopez Bernal J, Andrews N, Gower C, et al. Effectiveness of Covid-19 Vaccines against the B.1.617.2 (Delta) Variant. N Engl J Med 2021.

17. Sheikh A, McMenamin J, Taylor B, Robertson C. SARS-CoV-2 Delta VOC in Scotland: demographics, risk of hospital admission, and vaccine effectiveness. The Lancet 2021;397:2461–2.

18. Nasreen S, He S, Chung H, et al. Effectiveness of COVID-19 vaccines against variants of concern, Canada. medRxiv 2021:2021.06.28.21259420.

19. Al-Thani MH, Farag E, Bertollini R, et al. SARS-CoV-2 infection is at herd immunity in the majority segment of the population of Qatar. Open Forum Infectious Diseases 2021.

20. Jeremijenko A, Chemaitelly H, Ayoub HH, et al. Herd Immunity against Severe Acute Respiratory Syndrome Coronavirus 2 Infection in 10 Communities, Qatar. Emerg Infect Dis 2021;27:1343–52.

21. Coyle PV, Chemaitelly H, Kacem M, et al. SARS-CoV-2 seroprevalence in the urban population of Qatar: An analysis of antibody testing on a sample of 112,941 individuals. iScience 2021:102646.

22. World Health Organization. COVID-19 clinical management: living guidance. Available from: https://www.who.int/publications/i/item/WHO-2019-nCoV-clinical-2021-1. Accessed on: May 15 2021. 2021.

23. World Health Organization. International guidelines for certification and classification (coding) of COVID-19 as cause of death. Available from: https://www.who.int/classifications/icd/Guidelines_Cause_of_Death_COVID-19-20200420-EN.pdf?ua=1. Document Number: WHO/HQ/DDI/DNA/CAT. Accessed on May 31, 2021. 2021.

24. Multiplexed RT-qPCR to screen for SARS-COV-2 B.1.1.7, B.1.351, and P.1 variants of concern V.3. dx.doi.org/10.17504/protocols.io.br9vm966.2021. (Accessed June 6, 2021, at https://www.protocols.io/view/multiplexed-rt-qpcr-to-screen-for-sars-cov-2-b-1-1-br9vm966.)

25. Jacoby P, Kelly H. Is it necessary to adjust for calendar time in a test negative design?: Responding to: Jackson ML, Nelson JC. The test negative design for estimating influenza vaccine effectiveness. Vaccine 2013;31(April (17)):2165–8. Vaccine 2014;32:2942.

26. Pearce N. Analysis of matched case-control studies. BMJ 2016;352:i969.

27. Rothman KJ, Greenland S, Lash TL. Modern epidemiology. 3rd ed. Philadelphia: Wolters Kluwer Health/Lippincott Williams & Wilkins; 2008.

28. StataCorp. Stata Statistical Software: Release 17. College Station, TX: StataCorp LLC. 2021.

29. Abu-Raddad LJ, Chemaitelly H, Yassine HM, et al. Pfizer-BioNTech mRNA BNT162b2 Covid-19 vaccine protection against variants of concern after one versus two doses. J Travel Med 2021.

30. Tang P, Hasan MR, Chemaitelly H, et al. BNT162b2 and mRNA-1273 COVID-19 vaccine effectiveness against the Delta (B.1.617.2) variant in Qatar. medRxiv 2021:2021.08.11.21261885.

31. Abu-Raddad LJ, Chemaitelly H, Ayoub HH, et al. Effect of vaccination and of prior infection on infectiousness of vaccine breakthrough infections and reinfections. medRxiv 2021:2021.07.28.21261086.

32. Makhoul M., Ayoub H.H., Chemaitelly H., et al. Epidemiological impact of SARS-CoV-2 vaccination: Mathematical modeling analyses. Vaccines 2020;8.

33. Usherwood T, LaJoie Z, Srivastava V. A model and predictions for COVID-19 considering population behavior and vaccination. Scientific Reports 2021;11:12051.

34. Andersson O, Campos-Mercade P, Meier AN, Wengström E. Anticipation of COVID-19 Vaccines Reduces Social Distancing. https://papers.ssrn.com/sol3/papers.cfm?abstract_id=3765329. SSRN 2021.

35. Israel A, Merzon E, Schäffer AA, et al. Elapsed time since BNT162b2 vaccine and risk of SARS-CoV-2 infection in a large cohort. medRxiv 2021:2021.08.03.21261496.

36. Thomas SJ, Moreira ED, Kitchin N, et al. Six Month Safety and Efficacy of the BNT162b2 mRNA COVID-19 Vaccine. medRxiv 2021:2021.07.28.21261159.

37. Mizrahi B, Lotan R, Kalkstein N, et al. Correlation of SARS-CoV-2 Breakthrough Infections to Time-from-vaccine; Preliminary Study. medRxiv 2021:2021.07.29.21261317.

38. Pouwels KB, Pritchard E, Matthews PC, et al. Impact of Delta on viral burden and vaccine effectiveness against new SARS-CoV-2 infections in the UK Preprint 2021;2021.

39. Israel’s Ministry of Health. COVID-19 vaccine effectiveness against the Delta variant. Israel’s Ministry of Health report. https://www.gov.il/BlobFolder/reports/vaccine-efficacy-safety-follow-up-committee/he/files_publications_corona_two-dose-vaccination-data.pdf. 2021.

40. Nanduri S, Pilishvili T, Derado G, et al. Effectiveness of Pfizer-BioNTech and Moderna Vaccines in Preventing SARS-CoV-2 Infection Among Nursing Home Residents Before and During Widespread Circulation of the SARS-CoV-2 B.1.617.2 (Delta) Variant — National Healthcare Safety Network, March 1–August 1, 2021. Morbidity and Mortality Weekly Report 2021.

41. Rosenberg ES, Holtgrave DR, Dorabawila V, et al. New COVID-19 Cases and Hospitalizations Among Adults, by Vaccination Status — New York, May 3–July 25, 2021. Morbidity and Mortality Weekly Report 2021.

42. Puranik A, Lenehan PJ, Silvert E, et al. Comparison of two highly-effective mRNA vaccines for COVID-19 during periods of Alpha and Delta variant prevalence. medRxiv 2021:2021.08.06.21261707.

43. Abu-Raddad LJ, Chemaitelly H, Coyle P, et al. SARS-CoV-2 antibody-positivity protects against reinfection for at least seven months with 95% efficacy. EClinicalMedicine 2021;35:100861.

44. Abu-Raddad LJ, Chemaitelly H, Malek JA, et al. Assessment of the risk of SARS-CoV-2 reinfection in an intense re-exposure setting. Clin Infect Dis 2020.

## References

1. World Health Organization. COVID-19 clinical management: living guidance. Available from: https://www.who.int/publications/i/item/WHO-2019-nCoV-clinical-2021-1. Accessed on: May 15 2021. 2021.

2. World Health Organization. International guidelines for certification and classification (coding) of COVID-19 as cause of death. Available from: https://www.who.int/classifications/icd/Guidelines_Cause_of_Death_COVID-19-20200420-EN.pdf?ua=1. Document Number: WHO/HQ/DDI/DNA/CAT. Accessed on May 31, 2021. 2021.

3. Multiplexed RT-qPCR to screen for SARS-COV-2 B.1.1.7, B.1.351, and P.1 variants of concern V.3. dx.doi.org/10.17504/protocols.io.br9vm966. 2021. (Accessed June 6, 2021, at https://www.protocols.io/view/multiplexed-rt-qpcr-to-screen-for-sars-cov-2-b-1-1-br9vm966.)

4. Abu-Raddad LJ, Chemaitelly H, Butt AA, National Study Group for Covid-19 Vaccination. Effectiveness of the BNT162b2 Covid-19 Vaccine against the B.1.1.7 and B.1.351 Variants. N Engl J Med 2021.

5. Chemaitelly H, Yassine HM, Benslimane FM, et al. mRNA-1273 COVID-19 vaccine effectiveness against the B.1.1.7 and B.1.351 variants and severe COVID-19 disease in Qatar. Nat Med 2021.

6. Qatar viral genome sequencing data. Data on randomly collected samples. https://www.gisaid.org/phylodynamics/global/nextstrain/. 2021. at https://www.gisaid.org/phylodynamics/global/nextstrain/.)

7. Benslimane FM, Al Khatib HA, Al-Jamal O, et al. One year of SARS-CoV-2: Genomic characterization of COVID-19 outbreak in Qatar. medRxiv 2021:2021.05.19.21257433.

8. Hasan MR, Kalikiri MKR, Mirza F, et al. Real-Time SARS-CoV-2 Genotyping by High-Throughput Multiplex PCR Reveals the Epidemiology of the Variants of Concern in Qatar. medRxiv 2021:2021.07.18.21260718.

9. World Health Organization. Tracking SARS-CoV-2 variants. Available from: https://www.who.int/en/activities/tracking-SARS-CoV-2-variants/. Accessed on: June 5, 2021. 2021.

10. Abu-Raddad LJ, Chemaitelly H, Coyle P, et al. SARS-CoV-2 antibody-positivity protects against reinfection for at least seven months with 95% efficacy. EClinicalMedicine 2021;35:100861.

11. Abu-Raddad LJ, Chemaitelly H, Malek JA, et al. Assessment of the risk of SARS-CoV-2 reinfection in an intense re-exposure setting. Clin Infect Dis 2020.

12. Jackson ML, Nelson JC. The test-negative design for estimating influenza vaccine effectiveness. Vaccine 2013;31:2165–8.

13. Verani JR, Baqui AH, Broome CV, et al. Case-control vaccine effectiveness studies: Preparation, design, and enrollment of cases and controls. Vaccine 2017;35:3295–302.

